# THE TIME TO OFFER TREATMENTS FOR COVID-19

**DOI:** 10.1101/2020.05.27.20115238

**Authors:** Binh T. Ngo, Paul Marik, Pierre Kory, Leland Shapiro, Raphael Thomadsen, Jose Iglesias, Stephen Ditmore, Marc Rendell, Joseph Varon, Michael Dubé, Neha Nanda, Gino In, Daniel Arkfeld, Preet Chaudhary, Vito M. Campese, Diana L. Hanna, David E. Sawcer, Glenn Ehresmann, David Peng, Miroslaw Smogorewski, April Armstrong, Rajkumar Dasgupta, Fred Sattler, Denise Brennan-Rieder, Cristina Mussini, Oriol Mitja, Vicente Soriano, Nicolas Peschanski, Gilles Hayem, Marco Confalonieri, Maria Carmela Piccirillo, Antonio Lobo-Ferreira, Iraldo Bello Rivero, Eivind H. Vinjevoll, Daniel Griffin, Ivan FN Hung

**Affiliations:** Eastern Virginia Medical School, Department of Internal Medicine, Chief, Pulmonary and Critical Care Medicine, 825 Fairfax Ave, Rm 575, Norfolk, VA 23507; Pulmonary and Critical Care Medicine Aurora St. Luke’s Medical Center Milwaukee, Wisconsin.; Rocky Mountain Regional Veterans Affairs Medical Center in Aurora, CO and University of Colorado Anschutz Medical Campus in Aurora, CO Supported by The Emily Foundation in Boston, MA; Washington University in St. Louis; Jersey Shore University Medical Center, Neptune NJ, Hackensack Meridian School of Medicine at Seton Hall; Parkchester Times; The Rose Salter Medical Research Foundation Newport Coast, CA 92657; United Memorial Medical Center, University of Texas School of Medicine Houston, Texas, USA; CoronaTracker Community Research Group; University of Modena and Reggio Emilia in Italy; Hospital Universitari Germans Trias i Pujol Badalona, Spain; UNIR Health Sciences School & Medical Center C/ Almansa 101 Madrid 28040, Spain; UniversityHospital of Rennes Rennes, France; Hôpital Paris Saint-. Joseph, 75014 Paris, France.; Azienda Ospedaliero-Universitaria di Trieste Trieste, Italia; Istituto Nazionale Tumori, IRCCS, Fondazione G. Pascale Napoli, Italia; Unidade de Investigação Cardiovascular (UniC), Faculdade de Medicina da Universidade do Porto, Centro Hospitalar Universitário de São João, Porto, and Hospital Rainha Santa Isabel, Marco de Canaveses, Portugal; Department of Clinical Investigations, Center for Genetic Engineering and Biotechnology, Ave 31 e/ 158 and 190. Cubanacan, Playa, 10600 Havana, Cuba.; Volda Hospital HMR, Norway.; ProHEALTH, an OPTUM Company Columbia University, College of Physicians and Surgeons; Li Ka Shing Faculty of Medicine University of Hong Kong

## Abstract

**BACKGROUND:** As of January 1, 2021, there have been 81,947,503 confirmed cases of COVID-19, resulting in 1,808,041 deaths worldwide. Several vaccines are now available for emergency use, but it will take many months to immunize the world’s population. There is a pressing need for outpatient treatment now. We reviewed the possible options

**METHODS:** We reviewed up-to-date information from several sources to identify potential treatments to be utilized now for COVID-19. We searched for all ongoing, completed and published trial results with subject numbers of 100 or more, and used a targeted search to find announcements of unpublished trial results.

**RESULTS:** As of December 27, 2020, we identified 750 trials currently in recruitment phase. Of these, 122 were directed at prevention in healthy individuals, 100 were classified as treatment of outpatients with documented infection, and 390 were for treatment of hospitalized inpatients. There were 9 trials focusing on the post discharge Tail phase. Among the trials, there were 60 vaccine trials, 120 trials of hydroxychloroquine, 33 trials of alternative therapy, 12 trials of colchicine, 38 trials of anticoagulants, 22 trials of the RNA polymerase inhibitor favipiravir (FVP), 19 trials of interferons, 18 trials of glucocorticoid, and 58 trials of plasma based products. Closure of enrollment was projected by the end of the year for 153 trials. We found 83 publications reporting findings in human studies on 14 classes of agents, and on 7 vaccines. There were 45 randomized or active controlled studies, the rest retrospective observational analyses. Most publications dealt with hospitalized patients, only 18 publications in outpatients. Remdesivir, convalescent plasma, and synthetic anti-spike protein antibodies have been granted emergency use authorization in the United States. There is also support for glucocorticoid treatment of the COVID-19 respiratory distress syndrome. There is data supporting the use of several antiviral medications, some of which are in use in other countries.

**CONCLUSION:** Vaccines and antibodies are highly antigen specific. There is a need for antiviral agents in addition to mass immunization. It will be necessary for public health authorities to make hard decisions, with limited data, to prevent the continued spread of the disease and deaths.

## 1. INTRODUCTION

SARS-Cov-2 spread with extraordinary speed from its origin in December 2019 in Wuhan, China. As of January 1, 2021, there have been 81,947,503 confirmed cases of COVID-19, resulting in 1,808,041 deaths worldwide affecting 220 countries and territories (1). Symptomatic COVID-19 exhibits a characteristic sequence of phases related to the primary viral attack, manifesting as an influenza-like illness, affecting 80% of infected individuals. Then, in patients destined for severe disease, within seven to 10 days of onset of symptoms, an organizing pneumonia develops which requires assisted mechanical ventilation in up to 5% of patients (See Figure 1) (2, 3). Elevated cytokines and a coagulopathy at this pulmonary stage suggests an immune reaction as probable cause (4, 5).

**Figure 1:**
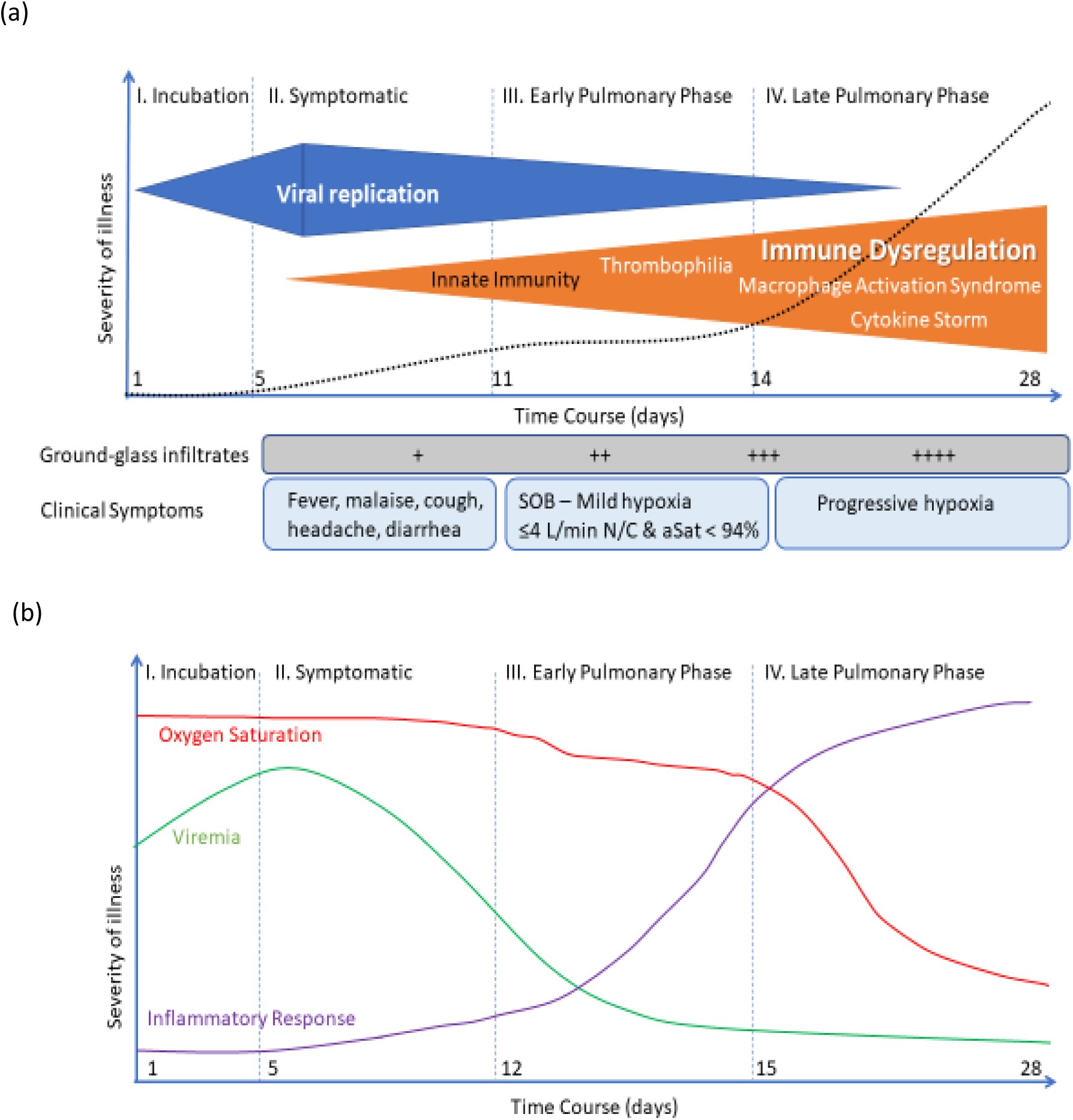
THE PHASES OF COVID-19 INFECTION. Asymptomatic viremia typically lasts for 5 days followed by symptoms of an upper respiratory infection. The immune response takes hold over the next week. Pneumonic infiltrates begin to appear. In patients destined for severe infection, a hyperinflammatory reaction takes hold, leading to worsening of pulmonary symptoms, hypoxia, and potentially acute respiratory distress syndrome.

In mainland China, vigorous efforts to contain the spread of COVID-19 by home isolation, closure of business activity, travel bans, and tracking and control of contacts succeeded. As of December 10, China has had 96,762 confirmed cases and 4,789 deaths, but the number of new cases has been low since April (1). Similar measures contained the epidemic in Taiwan and in New Zealand (1). However, the disease has not been controlled in the rest of the World, particularly Europe where there have been 26,490,355 cases with 582,016 deaths and the United States (USA) where, despite only a fifth of the population of China, there have been 19,746,390 confirmed cases of COVID-19 and 335,789 deaths. (1). In the Western World, as in China, the response to the pandemic was through reduction of person-to-person contact. Beginning in early March, social gatherings and meetings ended; schools and businesses were closed along with recreation areas, parks, and beaches. These social distancing efforts blunted the spread of the disease, but not with the results achieved in China. Due to economic pressures, epidemiologic containment was relaxed resulting in a significant ongoing surge well surpassing the original April peak.

The major industrialized countries have made an unprecedented effort to rapidly develop vaccines for COVID-19. Two mRNA vaccines have demonstrated 90% effectiveness at two months post immunization, and mass vaccination has begun. However, in view of the continuing daily increase in new cases and deaths, we believe that it is unwise to place sole reliance on immunization alone. Our goal was to assess the options at the present time for release of pharmacologic agents to treat and to prevent COVID-19 while pursuing widespread vaccination in 2021.

## 2. METHODS

### 2.1 CLINICAL TRIALS

We reviewed up-to-date information from multiple sources to identify potential treatments for COVID-19: The Reagan-Udall Expanded Access Navigator COVID-19 Treatment Hub was used to track the efforts of companies to develop therapeutic interventions. We searched for investigational trials in active recruitment and those that have completed enrollment. We used (A) covid-trials.org, a registry to collate all trials in real time with data pulled from the International Clinical Trials Registry Platform and all major national registries (6). We cross validated information on (B) clinicaltrials.gov, the registry of clinical trials information maintained by the United States National Library of Medicine. We further cross referenced the trials on C) the World Health Organization’s International Clinical Trials Registry Platform (WHO ICTRP), and (D) (Cochrane COVID-19 Study Register. We excluded studies which were clearly observational with multiple different treatments, with no means of comparison. We further set a cutoff of 100 for numbers of subjects since smaller trial size typically lacks statistical power to enable regulatory approval. For each trial selected, we documented the setting of patient contact, either hospital or outpatient, the type of control procedure, the date the trial was initially registered, and the proposed date of completion of enrollment.

### 2.2 PUBLISHED TRIAL RESULTS

Our search was carried on the week of December 27, 2020. We identified all publications on pubmed.gov to find peer reviewed articles, on medRxiv to find preprint reports and the WHO Global Literature on Coronavirus Disease. We further carried out daily Google™ searches on each potential treatment to find preliminary reports, typically presented as press releases, reviewed by a journalist. We included all publications in our results as well as trial results posted as complete on Clinicaltrials.gov. We did not exclude trials with less than 100 subjects, since many reports were interim, with trials ongoing.

### 2.3 VIRTUAL DISCUSSIONS AMONG CO-AUTHORS

We used the preprint server medRxiv to post a systematic analysis of the development of therapeutic interventions for COVID-19 in order to stimulate diffusion of the manuscript and allow widespread “open source” input from co-authors (7). Successive versions of the preprint article have been posted periodically as a chronicle beginning in late May, 2020 and continuing to date (8). This process led to the present multinational consensus process involving 36 co-authors.

## 3. RESULTS

### 3.1 REVIEW OF CURRENT TRIALS

As of December 27, 2020, we identified 750 trials currently in recruitment phase with subject size of 100 or more (Supplementary Table 1). Of these, 122 were directed at prevention in healthy individuals, 100 were classified as treatment of outpatients with documented infection, and 390 were for treatment of hospitalized inpatients. There were 9 trials focusing on the post discharge Tail phase. The remaining trials were unclear as to intended subjects. Among the trials, there were 60 vaccine trials, 120 trials of hydroxychloroquine (HCQ), 33 trials of alternative therapy, 12 trials of colchicine, 38 trials of anticoagulants, 22 trials of the RNA polymerase inhibitor favipiravir (FVP), 19 trials of interferons, 18 trials of glucocorticoid, and 58 trials of plasma based products. Closure of enrollment was projected by the end of the year for 153 trials.

### 3.2 COMPLETED TRIALS

As of December 27, 2020, there were 97 trials reporting completion with 100 or more subjects(Supplementary Table 2 There were 52 trials in hospitalized patients, 9 directed at outpatients, and 12 prevention studies, the rest mixed.

### 3.3 PUBLISHED RESULTS ON COVID-19 TRIALS

As of December 27, 2020 we found 83 publications reporting findings in human studies on 14 classes of agents, and on 7 vaccines (See TABLE 1). There were 45 randomized or active controlled studies. The rest were retrospective observational analyses. Only 15 publications dealt with outpatient care, including 7 vaccine reports; the rest were all in hospitalized patients.

**TABLE 1.**
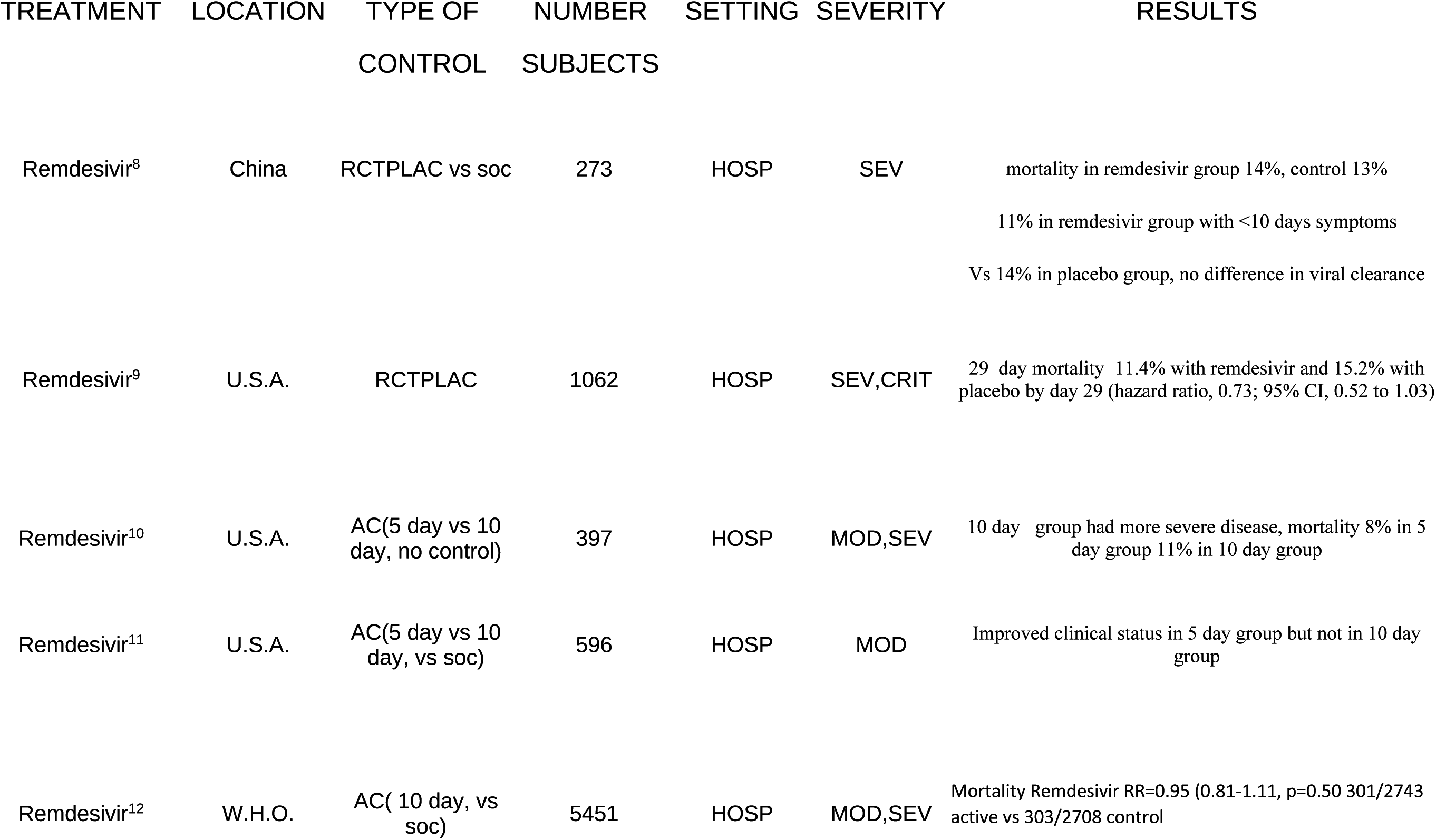
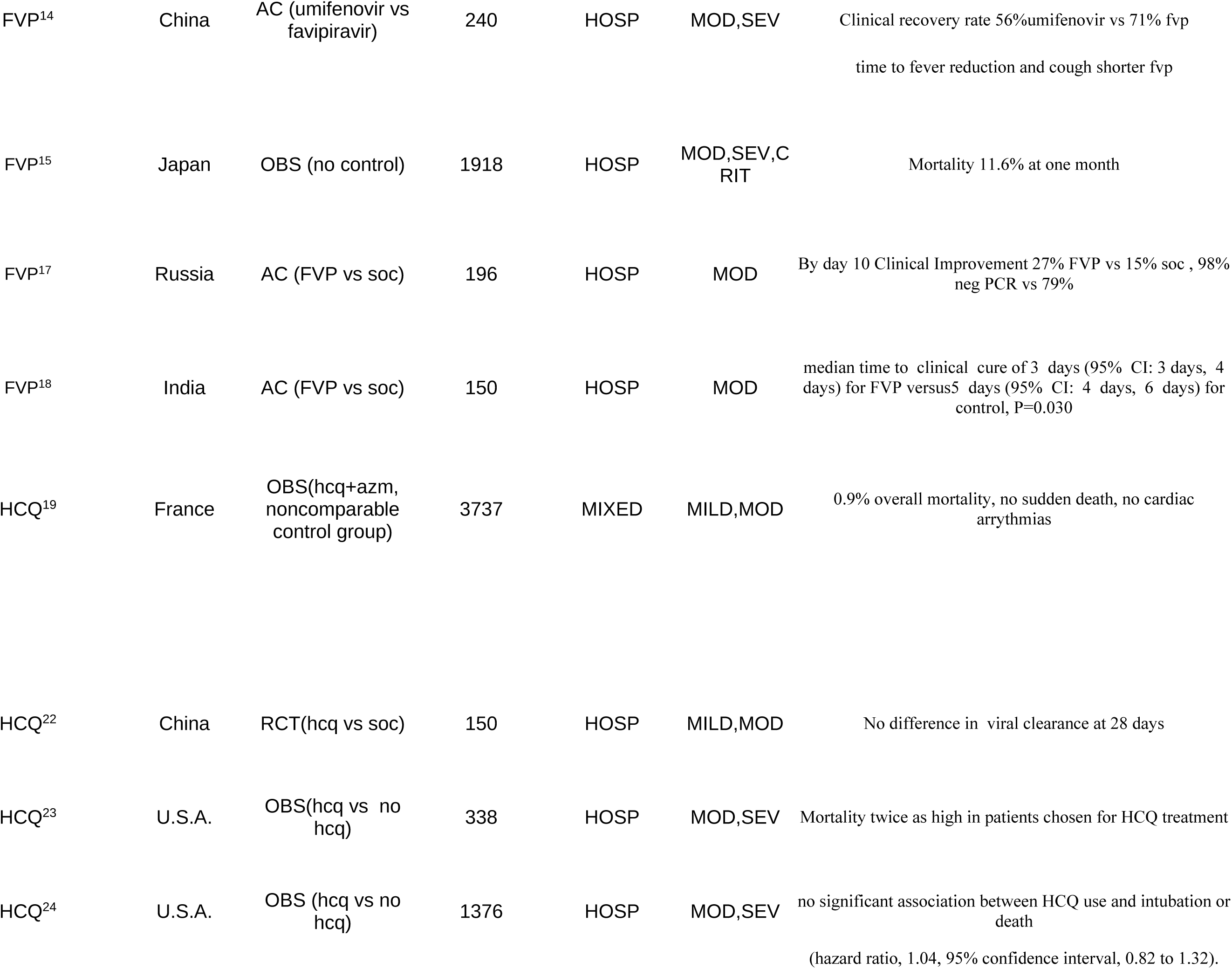
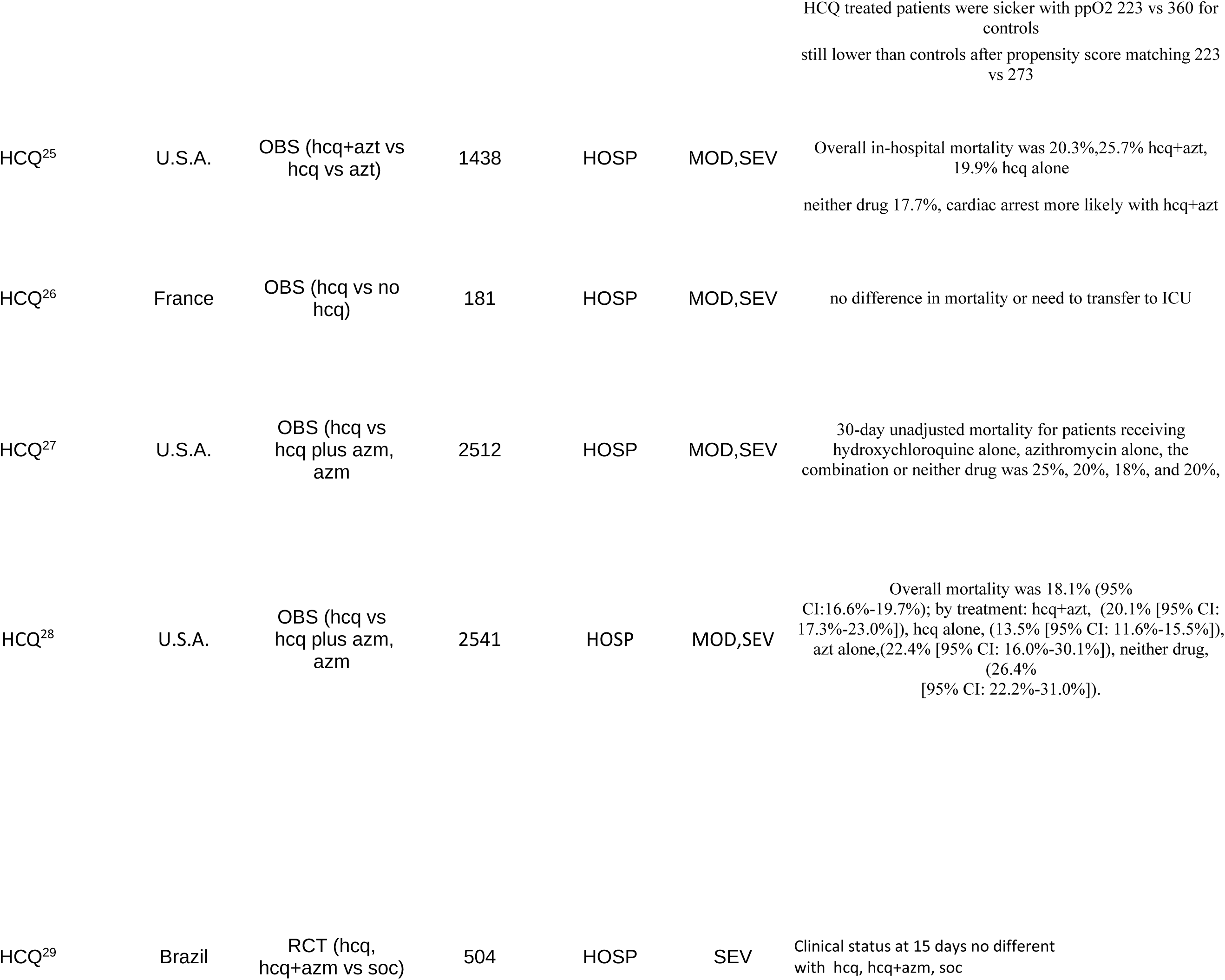
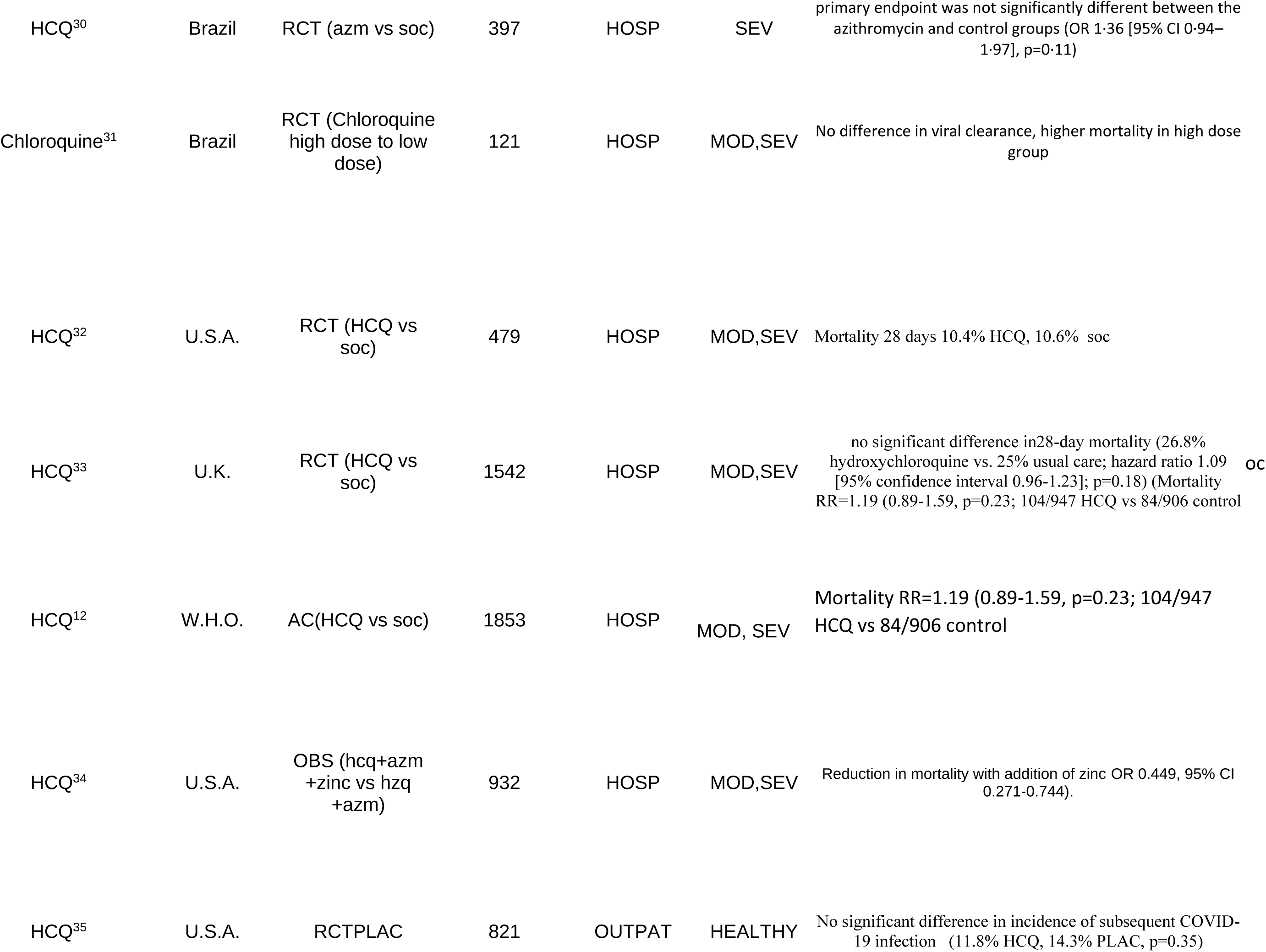
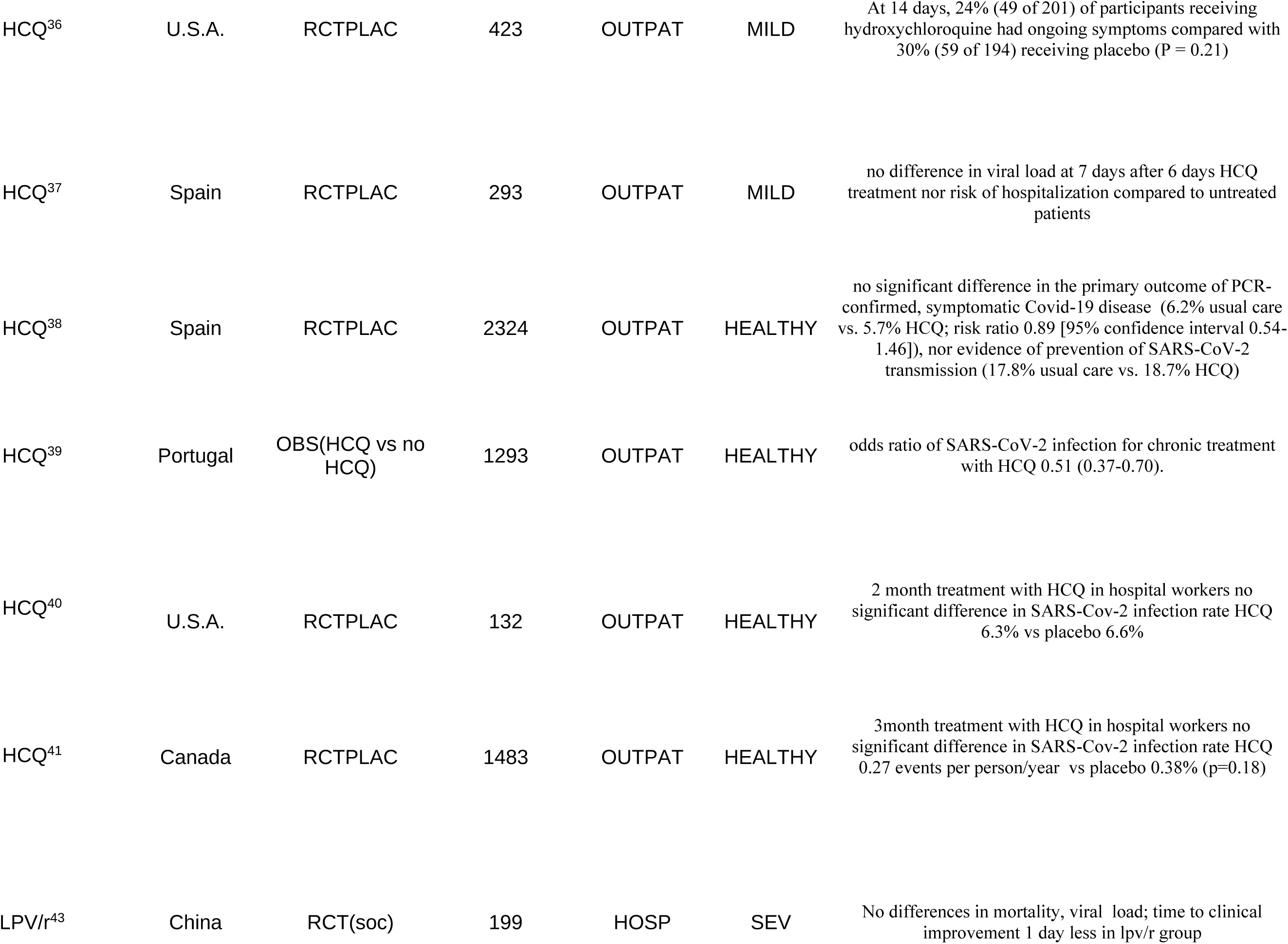
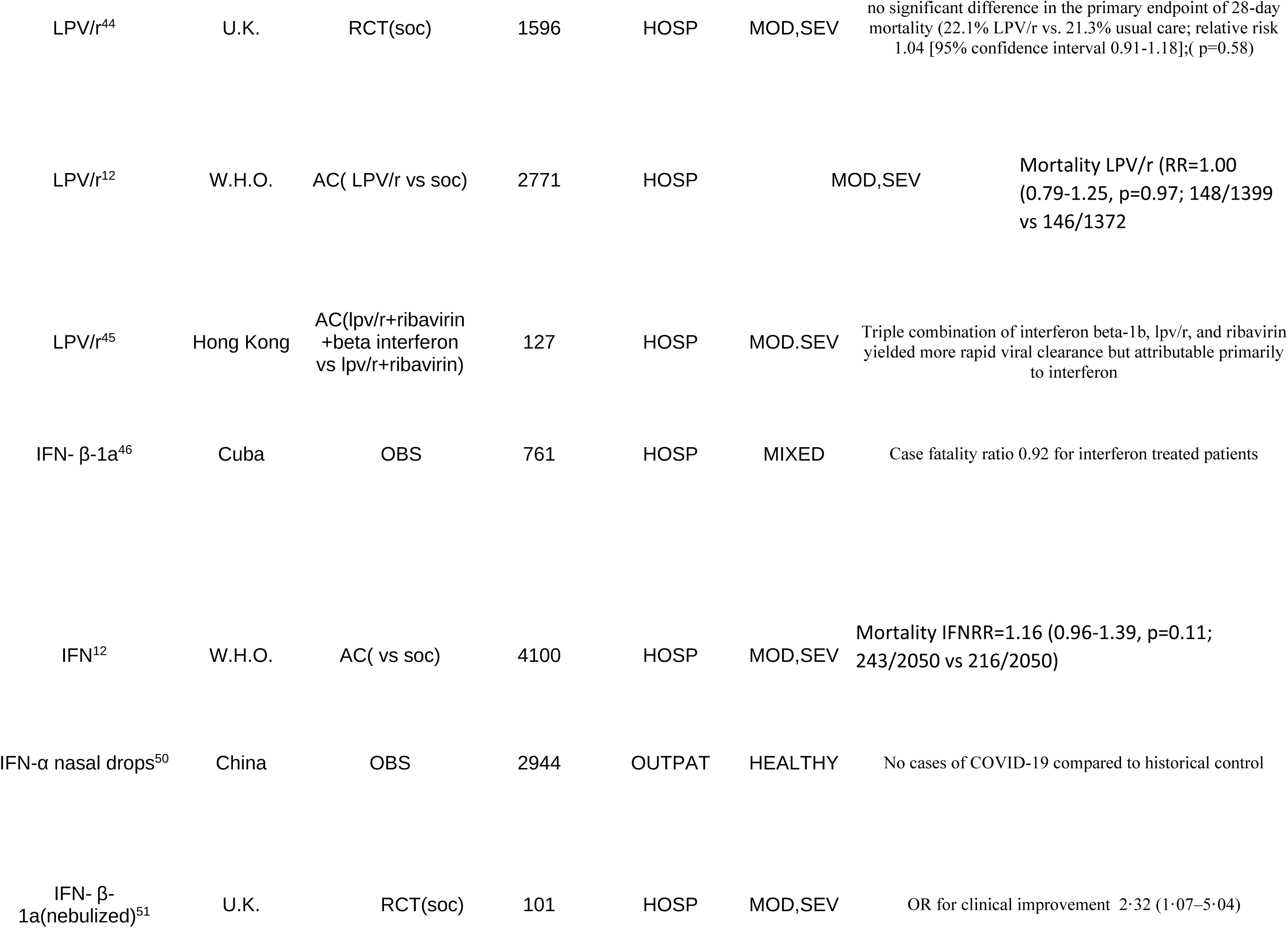
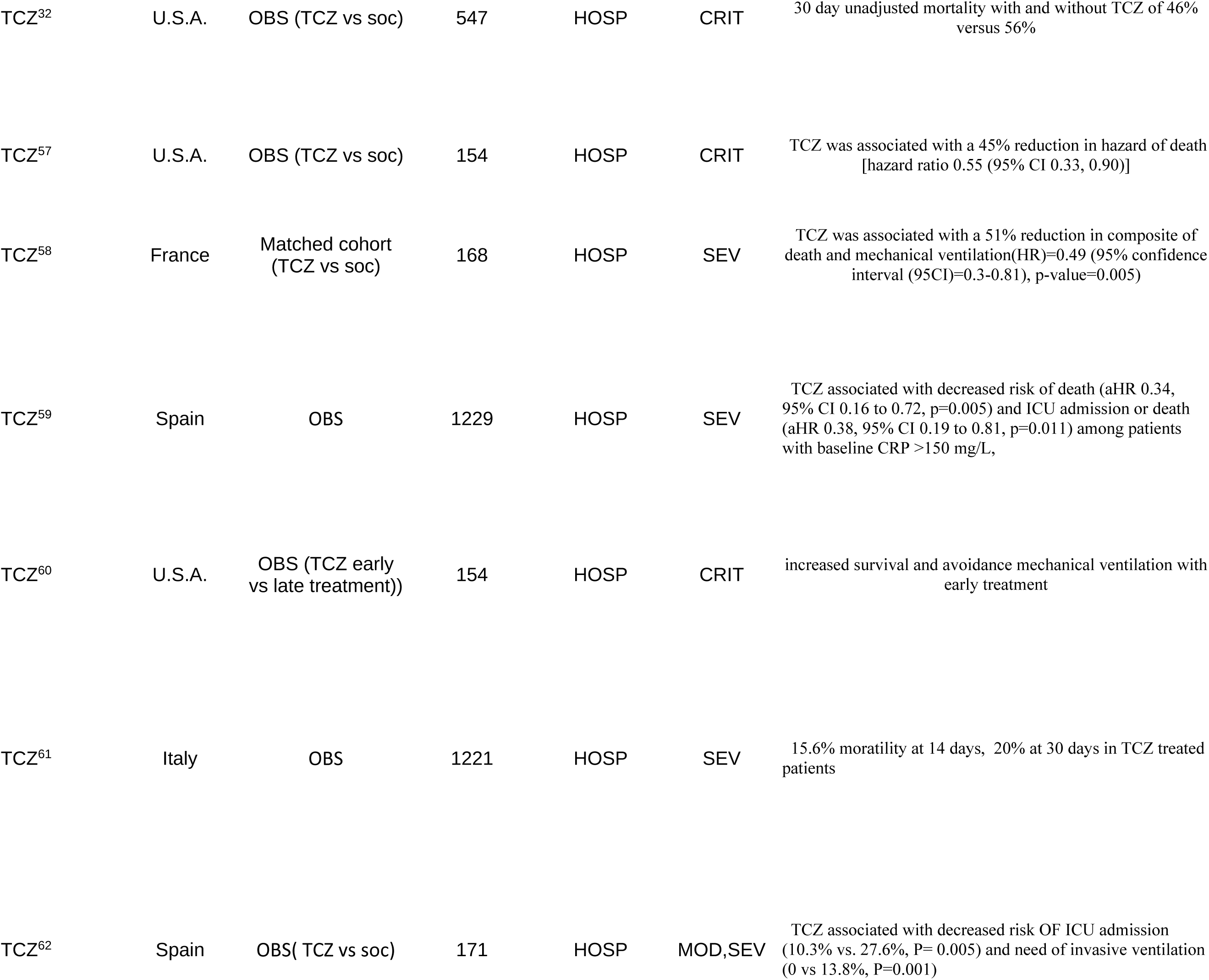
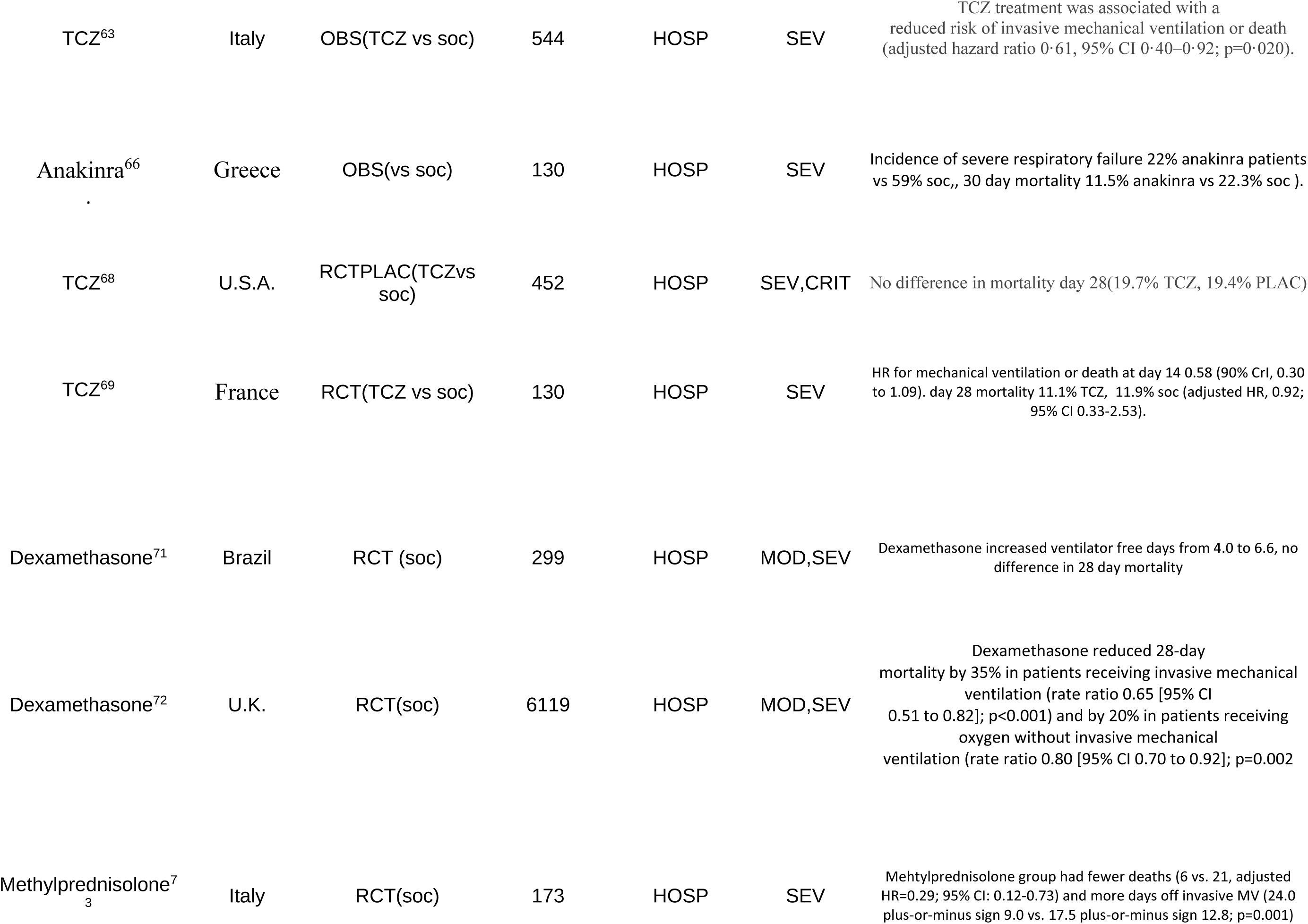
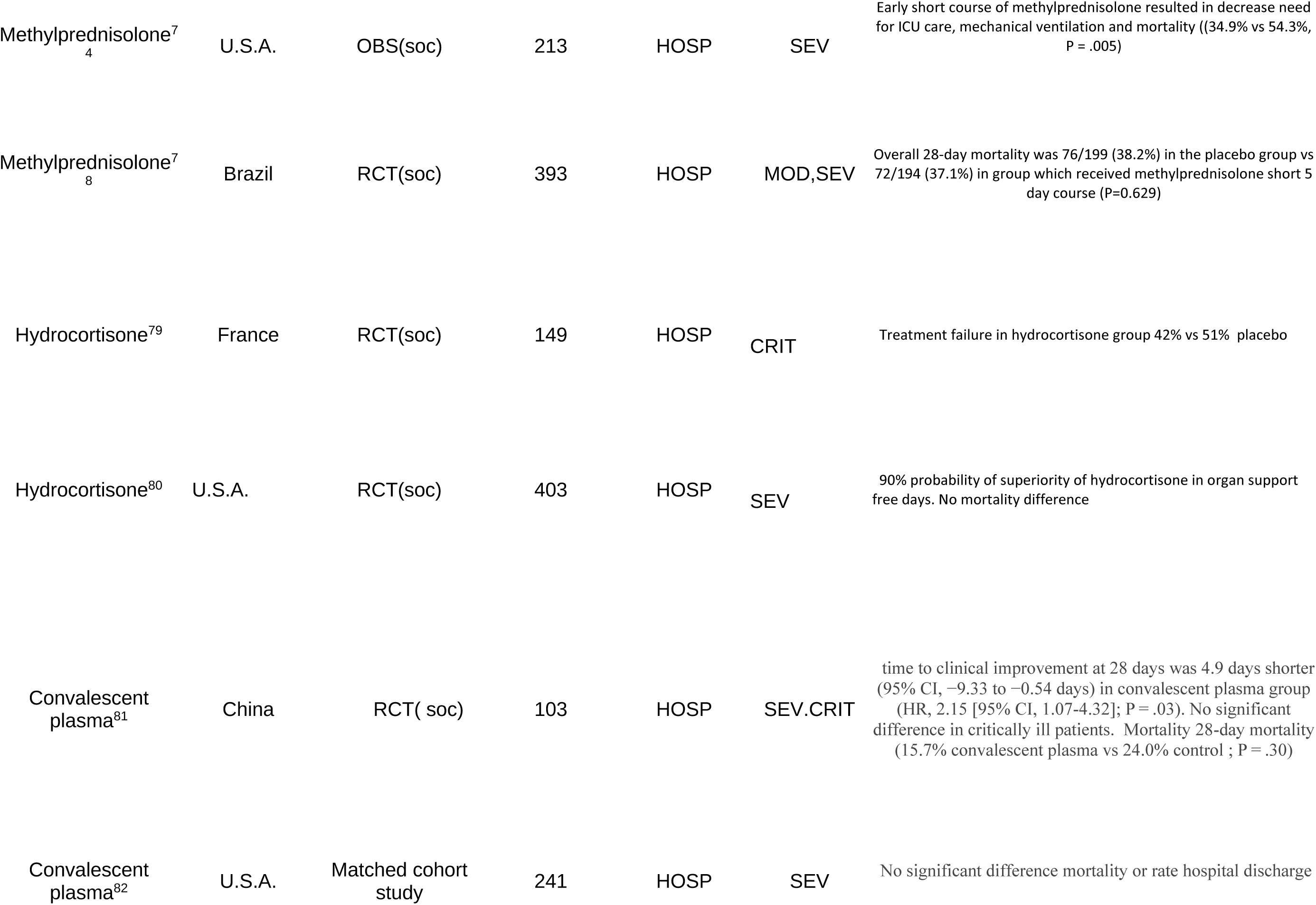
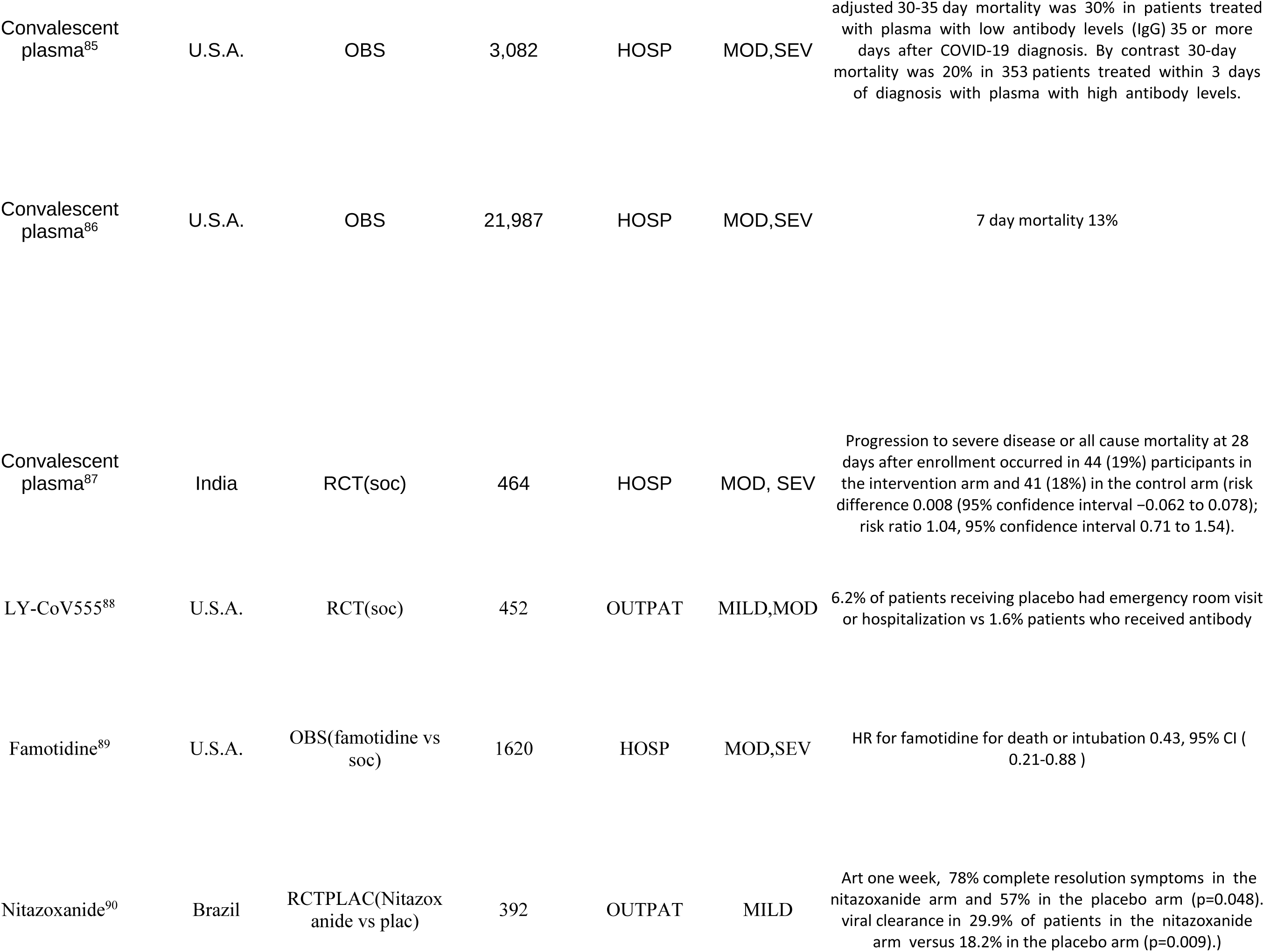
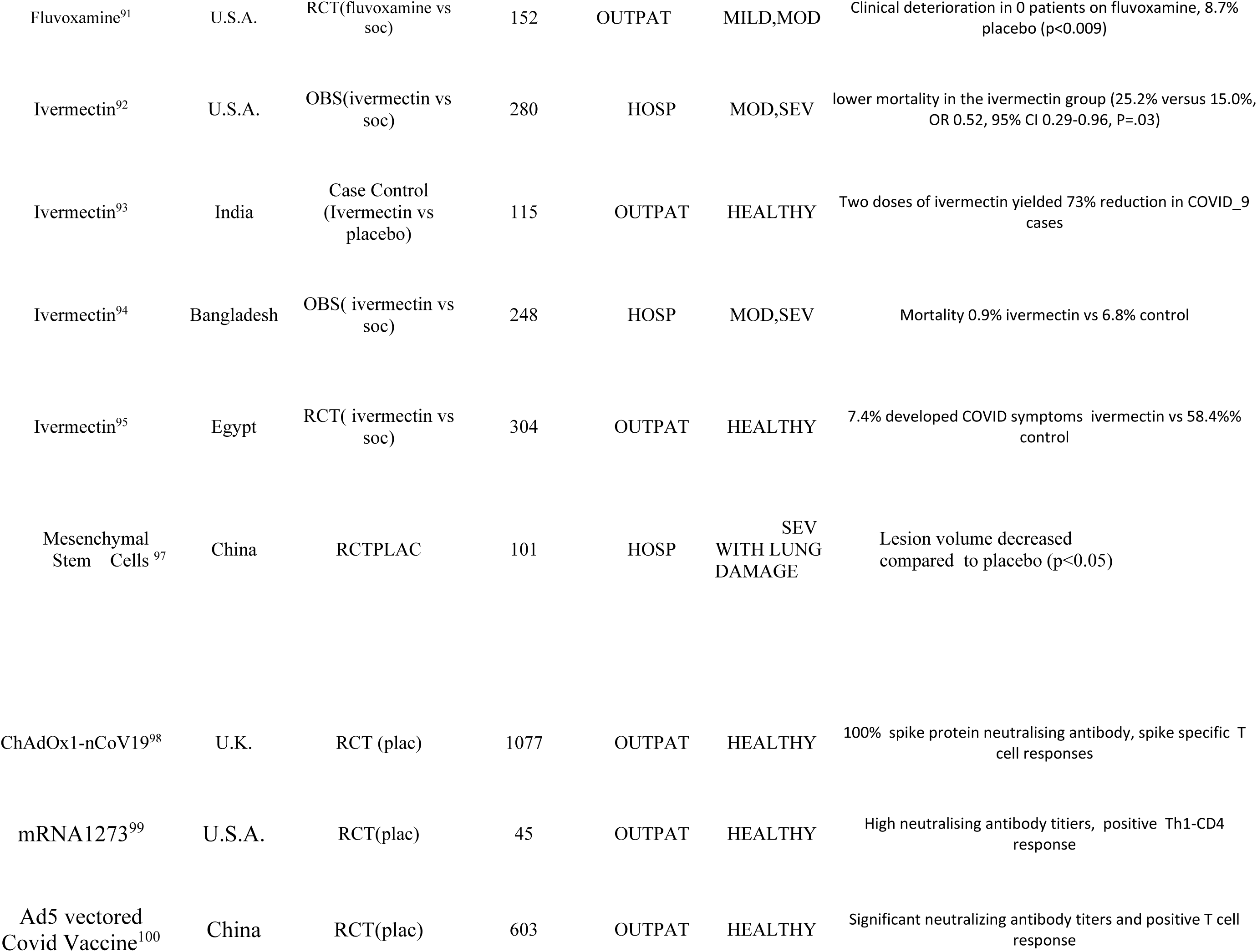
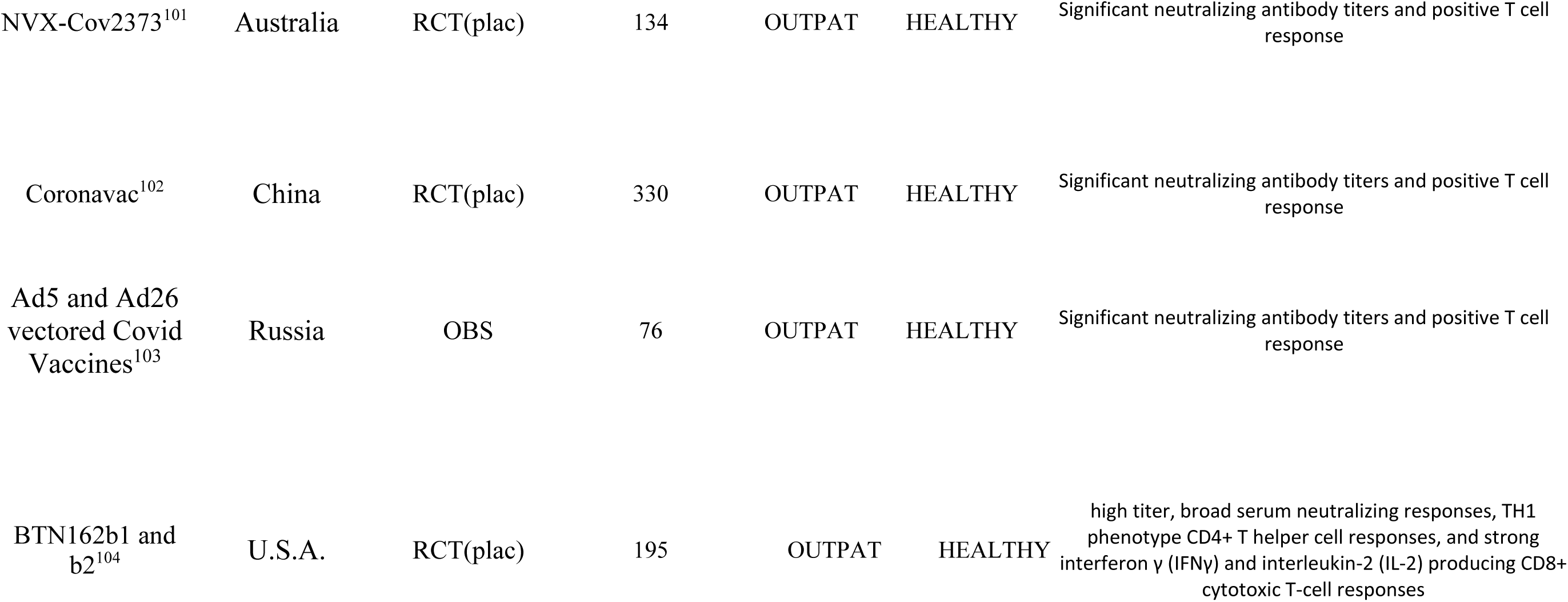
PUBLISHED STUDIES AS OF DECEMBER 27, 2020. Each agent is listed along with the country originating the publication. We have listed studies with 100 or more subjects. The type of control procedure: RCT: Randomized controlled study PLAC: Placebo, soc: standard of care which is variable depending on each location. AC: Active control: the control options are listed in parenthesis. OBS: retrospective observation study. LPV/R: lopanovir/ritonavir; HCQ: hydroxychloroquine; AZM: azithromycin; IFN: interferon; tocilizumab: TCZ. SEV: severe; CRIT: critical; MOD: moderate. W.H.O.: World Health Organization

**3.3.1** In the group of antiviral agents, the largest published randomized, controlled trials were with the intravenously administered RNA polymerase inhibitor remdesivir (9–13). A double blind, randomized placebo controlled trial of remdesivir, in 1062 moderate to severely ill patients was carried out by the National Institute of Allergy and Infectious Disease (NIAID) (10). The patients who received remdesivir had a median recovery time of 10 days (95% confidence interval [CI], 9 to 11), as compared with 15 days (95% CI, 13 to 18) among those who received placebo (rate ratio for recovery, 1.29; 95% CI, 1.12 to 1.49; P<0.001). P<0.001). The Kaplan-Meier estimates of mortality by 29 days after randomization were 11.4% with remdesivir and 15.2% with placebo (hazard ratio, 0.73; 95% CI, 0.52 to 1.03). On the basis of this trial, the Food and Drug Administration (FDA) approved the use of remdesivir for COVID-19 hospitalized patients. A different study of remdesivir in hospitalized patients with moderate COVID-19 clinical status showed improved clinical status by day 11 compared to standard of care for a 5 day course of remdesivir, but not for a 10 day course (12). There has been no data provided on viral clearance for the U.S. remdesivir studies. In early April, the WHO organized a megatrial, appropriately named Solidarity, to assess four separate treatment options only in hospitalized patients: (a) Remdesivir, an intravenously administered inhibitor of RNA polymerase; (b) The HIV agent lopanovir/ritanovir (LPV/R); (c) LPV/R plus Interferon β-1a; and (d) the antimalarials hydroxychloroquine (HCQ) and chloroquine (13). Results of the Solidarity Trial were reported by the World Health Organization on October 15, 2020. They found no effect of remdesivir on 28 day mortality, need for mechanical ventilation nor duration of hospitalization in a study including 5451 hospitalized patients (13).

**3.3.2** There were two positive randomized, active control clinical trials in China of the orally administered RNA polymerase inhibitor favipiravir (FVP) (14, 15). In a trial comparing 116 patients on FVP to 120 on umifenovir, the 7 day clinical recovery rate was 55.9% for the umifenovir group and 71% in the FVP group (15). In Russia, a preliminary study of FVP showed reduced duration of viral shedding, and the drug was approved for clinical treatment beginning in multiple hospitals on June 12, 2020 (17). In a follow-up phase 3 study, they reported 27% clinical improvement at day 10 compared to 15% for standard care with 98% clearance of SARS-COV-2 compared to 79% (18). In India, a study of 150 mild to moderate COVID-19 patients showed median time to clinical cure of 3 days (95% CI: 3 days, 4 days) for FVP versus 5 days (95% CI: 4 days, 6 days) for control, P=0.030, and FVP was approved to treat COVID-19 in July (19). In late September, a report, still unpublished, of a randomized controlled study in Japan announced more rapid viral clearance in FVP treated patients.

**3.3.3** There have been many studies of HCQ (20–47). A group in Marseille reported their experience with 3737 patients screened positive for SARS-Cov-2 and immediately treated with HCQ and azithromycin (AZM), after excluding patients at risk of QT prolongation (20). They showed clearance of viral shedding in 89.4% of the patients by ten days. The overall mortality in their population was 0.9%, none in patients under 60 years old, and no sudden cardiac deaths. They had no randomized control population, but their study prompted immediate widespread use of HCQ throughout the world for treatment of COVID-19 patients.

Subsequent randomized controlled studies in hospitalized patients have shown no clinical nor mortality benefit of HCQ. In the randomized RECOVERY trial in 1542 hospitalized patients in the United Kingdom, there was no significant difference in the primary endpoint of 28-day mortality (26.8% HCQ vs. 25% usual care; hazard ratio 1.09 [95% confidence interval 0.96-1.23]; *p*=0.18) (34). In the Solidarity Trial, there was no benefit of HCQ on mortality (HCQ RR=1.19 (0.89-1.59, p=0.23; 104/947 vs 84/906), need for mechanical ventilation, nor duration of hospitalization (13). However, in a retrospective comparison study at NYU Langone Health for all patients admitted between March 2 and April 5, 2020, there was a move to add zinc 100 mg daily to their standard HCQ plus AZM regimen (35). There was a significantly lower mortality (13.1%) among the zinc treated patients compared to those who did not receive zinc (22.8%).

There has been limited study of HCQ in outpatients. In one internet-based prevention study, with HCQ given for 5 days to healthy individuals with a significant exposure to SARS-Cov-2, the incidence of new illness compatible with COVID-19 did not differ significantly between participants receiving HCQ (49 of 414 [11.8%]) and those receiving placebo (58 of 407 [14.3%]); absolute difference −2.4 percentage points (95% confidence interval, −7.0 to 2.2; *p*=0.35) (36). The same group also treated 423 patients with mild symptoms imputed to COVID-19 (37). At 14 days, 24% (49 of 201) of participants receiving HCQ had ongoing symptoms compared with 30% (59 of 194) receiving placebo (*p* = 0.21).

A study in Spain of 293 patients with PCR-confirmed mild COVID-19, found no difference in viral load nor risk of hospitalization following 6 days of HCQ treatment compared to untreated patients (38). The same group treated 1,116 healthy contacts of 672 Covid-19 index cases with HCQ while 1,198 were randomly allocated to usual care (39). There was no significant difference in the primary outcome of PCR-confirmed, symptomatic Covid-19 disease (6.2% usual care vs. 5.7% HCQ; risk ratio 0.89 [95% confidence interval 0.54-1.46]), nor evidence of prevention of SARS-CoV-2 transmission (17.8% usual care vs. 18.7% HCQ).

A large population study in Portugal surveyed all patients on chronic HCQ treatment and cross-verified against a mandatory database of patients registered with COVID-19 (40). The incidence of a positive PCR test for SARS-CoV-2 infection in patients receiving HCQ was 5.96%, compared to 7.45% in those not so treated, adjusted odds ratio 0.51 (0.37-0.70). However, two separate randomized controlled studies found no evidence of benefit of two months prophylactic treatment of hospital workers with HCQ (41, 42).

**3.3.4**. Published results with Lopanovir/ritonavir (LPV/r) have been disappointing (13, 43–45). In a randomized trial with 99 patients on LPV/r and 100 receiving standard care, there was no difference in time to clinical improvement, mortality nor viral clearance (44). In the RECOVERY trial in 1,596 hospitalized patients there was no significant difference in the primary endpoint of 28-day mortality (22.1% LPV/r vs. 21.3% usual care; relative risk 1.04 [95% confidence interval 0.91-1.18]; (*p*=0.58) (45). In the Solidarity Study, there was no effect of LPV/r on 28 day mortality LPV/r RR=1.00 (0.79-1.25, p=0.97; 148/1399 vs 146/1372), need for mechanical ventilation nor duration of hospitalization (13).

A possibly more favorable result was obtained in Hong Kong in a study of 86 patients assigned to triple combination therapy with LPV/r plus ribavirin plus interferon β-1 (46). Compared to a control group of 41 patients receiving LPV/r alone, the combination group had a significantly shorter median time from start of study treatment to negative SARS-Cov-2 nasopharyngeal swab (7 days [IQR 5–11]) than the control group (12 days [8– 15]; hazard ratio 4.37 [95% CI 1.86–10.24], *p*=0·0010). The time to complete alleviation of symptoms was 4 days [IQR 3–8] in the combination group vs 8 days [7–9] in the control group; HR 3.92 [95% CI 1.66–9.23. This study was interpreted not as supporting LPV/r therapy, but rather focusing on interferon β-1 as the primary treatment modality.

**3.3.5** Interferon studies have been encouraging. In Cuba, a combination of interferon-α-2b and interferon γ on background therapy of LPV/r and chloroquine was successful in achieving viral clearance in 4 days in 78.6% of the patients compared to 40.6% of those receiving interferon-α-2b alone (49). In the Solidarity Study in hospitalized patients, there was no effect of injected interferon on 28 day mortality (IFN RR=1.16 (0.96-1.39, p=0.11; 243/2050 vs 216/2050), need for mechanical ventilation nor duration of hospitalization (13). However, a separate trial of nebulized interferon β-1 reported positive improvement in clinical status (52).

**3.3.6** There have been many observational studies of immunomodulatory therapy with the IL-6 inhibitor tocilizumab and the IL-1 inhibitor anakinra to mitigate the pulmonary phase of COVID-19 suggesting reduction of mortality and need for intubation in hospitalized severe and critically ill patients (28, 53–66). However, in several randomized control studies, there has been no benefit with tocilizumab (67–69). In the COVACTA trial, there was no benefit of tocilizumab in clinical status, mortality (19.7% tocilizumab vs placebo = 19.4%), nor in ventilator free days (68), and in the Cor-Immuno-Toci trial, no decrease in mortality at day 28 (69). The tocilizumab results in these randomized controlled trials were affected by a higher frequency of glucocorticoid use in the control groups.

**3.3.7** Pre-COVID-19, glucocorticoid treatment had been a recognized treatment with mortality benefit for patients in severe respiratory distress (70). Dexamethasone has now demonstrated benefit in several COVID-19 hospital trials (71–72). In the RECOVERY trial, 2104 patients were randomly allocated to receive dexamethasone for ten days compared to 4321 patients concurrently allocated to usual care (72). Overall, 454 (21.6%) patients randomized to dexamethasone and 1065 (24.6%) patients receiving usual care died within 28 days (age adjusted rate ratio [RR] 0.83; 95% confidence interval [CI] 0.74 to 0.92; *p*<0.001). Dexamethasone reduced deaths by one-third in the subgroup of patients receiving invasive mechanical ventilation (29.0% vs. 40.7%, RR 0.65 [95% CI 0.51 to 0.82]; *p*<0.001) and by one-fifth in patients receiving oxygen without invasive mechanical ventilation (21.5% vs. 25.0%, RR 0.80 [95% CI 0.70 to 0.92]; *p=*0.002), but did not reduce mortality in patients not receiving respiratory support at randomization (17.0% vs.13.2%, RR 1.22 [95% CI 0.93 to 1.61]; *p*=0.14). Most trials of other glucocorticoids have shown similar benefits in COVID-19 patients with acute respiratory failure (73–80)

**3.3.8** There is promising data on convalescent plasma (81–86). A meta-analysis including 4173 patient outcomes across a multinational series of studies concluded that convalescent plasma treatment yielded a 57% reduction in mortality (85). The reduction was greatest in patients treated within 3 days of diagnosis with high titer antibody plasma. Based on evidence of safety in over 20,000 patients in an expanded access program coordinated by the Mayo Clinic, the U.S. FDA recently issued an emergency authorization for use of convalescent plasma (86). Monoclonal antibodies to the SARS-COV-2 spike protein have been synthesized. Treatment of ambulatory patients with the LY-CoV555 neutralizing antibody and the Regeneron combination of two antibodies early in the course of infection has shown success in reducing the frequency of hospitalization, and the FDA issued an emergency authorization for use in outpatients (88).

**3.3.9** There have been a number of reports on agents not ordinarily considered as antivirals. These include famotidine, nitazoxanide, fluvoxamine, colchicine, and ivermectin (89–96). Colchicine, an anti-inflammatory agent, suggested benefit in the GRECCO, placebo-controlled, randomized clinical trial of 105 patients (90). The primary clinical end point of deterioration to mechanical ventilation or death was 14% in the control group (7 of 50 patients) and 1.8% in the colchicine group (1 of 55 patients) (odds ratio, 0.11; 95% CI, 0.01-0.96; p = 0.02). There have now been a number of trials including several randomized controlled studies suggesting benefit with ivermectin both in prevention and in treatment of COVID-19 (TABLE 1).

**3.3.10** A number of vaccines have reported successful induction of neutralizing antibody and cellular immune responses in early phase trials (98–104). The Moderna and Pfizer Biontech mRNA spike protein vaccines released data showing over 90% effectiveness at preventing symptomatic infection in a 2 month post immunization observation period, and emergency authorization for immunization has been given. Massive vaccinations have begun in many countries.

## 4. CONCLUSION

SARS-Cov-2 fueled by modern day jet travel overwhelmed medical preparedness for the pandemic. Current projections are for a continuation of new cases and deaths continuing in 2021 (105, 106). On December 31, 2020, 470,046 new cases were reported worldwide (1). Of these 174, 814 cases occurred in the United States. Clinical trials to date have focused primarily on hospitalized patients to try to prevent death. It is hoped that the use of remdesivir, convalescent plasma, glucocorticoids, anticoagulation and improved management of COVID-19 respiratory failure will lower the present rate of in hospital mortality which in some series has been as low as 6% but in most hospitals still approaches 20% (77). However, it is essential to change the present dynamic of attempted therapeutic responses to recognize specific phases of the disease (107). In particular, efforts must now focus on prevention and treatment of the initial viral infection so that hospitalization can be avoided. We face a dilemma with inadequate current prevention and treatment for outpatients with mild to moderate disease, constituting 80% of the infected population and the primary mode of spread of SARS-Cov-2. The lessons learned in very sick hospitalized patients do not necessarily apply to the earlier viremic phase. Antiviral agents, such as remdesivir and favipiravir, interferon, convalescent plasma, and monoclonal antibodies are likely to be most effective during the early stage of viremia, which is prior to the pulmonary phase requiring hospitalization.

Vaccines have been the solitary hope held out by public health authorities to arrest SARS-Cov-2. Progress on vaccine development has been rapid, but the speed of development does not reduce the challenges of developing vaccines to be administered to the population of the entire World. Although neutralizing antibodies and memory T cells can be produced by the vaccine candidate, the demonstration of true protection requires long-term follow-up of an exposed vaccinated population. The current vaccines will be released for emergency use after only a short two month period of observation. No matter how rapidly a vaccine advances to Phase III testing, the duration of follow-up cannot be shortened, and for SARS-Cov-2 possibly prolonged due to several factors. First, there is a risk of immune enhancement of infection which occurs when induced antibodies increase entry and internalization of virus into myeloid cells (108). This major complication struck the newly developed dengue vaccine in 2017 (109). Second, the sinister autoimmune pathogenicity of SARS-Cov-2 raises the risk of delayed long-term harm if the virus is not fully and immediately eradicated by the initial immune response to the vaccine. Furthermore, studies of recurrent coronavirus infections suggest that infection may confer immunity for less than a year (110). After implementation of a new vaccine, efforts to find volunteers for trials of new, more effective vaccines will be hampered if there is no treatment available for placebo treated patients who become infected. Vaccines are specific to the intended pathogen. There is the risk that the virus will mutate to a form for which the new vaccine may not be fully protective. Already two variants have emerged and caused major outbreaks. The level of protection offered by the current vaccines is currently under study. Finally, the degree of acceptance of the COVID-19 vaccines by the general population is questionable. Seasonal influenza vaccination coverage was only 48% in 2019 (111). Children typically have limited COVID-19 symptoms. However, they may serve as a reservoir of ongoing infectivity (112). Currently there have been no clinical trial results of vaccine in children.

It also follows that totally new viruses may emerge just as did SARS-Cov-2. COVID-19 is now the sixth severe viral epidemic to hit mankind in the past 20 years; certainly it has been more widespread than SARS-Cov-1, H1N1 influenza, MERS, Zika, and Ebola, but that does not diminish the gravity of these repeated viral threats. Development of antigen specific vaccines takes a long time during which disease takes a large toll as has been the case for SARS-Cov-2. The succession of viral afflictions points to the need to implement widespread use of antiviral agents. The current recom-mendation for COVID-19 is home quarantine with no specific treatment for patients with suspicious symptoms. What is needed is therapeutic intervention which can be used to treat all outpatients with positive COVID-19 tests at the time of initial symptoms, not waiting for deterioration requiring hospital care. Hydroxychloroquine has not succeeded. Remdesivir administered intravenously for five days is not a practical daily outpatient treatment; Gilead© is attempting to develop an inhaled remdesivir formulation, but those efforts are only now beginning. Favipiravir, a tablet, could be used in early stages of infection and has now been released in Russia, Hungary, and in India, but not in the United States and the European Union. It has known embryogenic risks, so its use requires restrictions on women of childbearing potential, as is the case with tretinoin for acne and thalidomide for multiple myeloma. Trials with camostat, the serine protease inhibitor, capable of blocking cell entry of SARS-Cov-2, are not due to end till the fourth quarter of the year. Similarly, a prevention trial with colchicine is not scheduled to end until the fourth quarter of 2020. Convalescent plasma is used primarily in hospital patients. Monoclonal antiviral neutralizing antibodies based on the same principle have finally received emergency use authorization for ambulatory patients. However, the scale of production of these monoclonal antibodies is far too limited to offer to all outpatients who should be treated. Interferon formulations have shown promise in several studies. There have been several randomized controlled studies suggesting benefit with ivermectin both for prophylaxis and treatment.

Yet, at this time, no outpatient therapy has been proven safe and effective in large scale phase III randomized, placebo-controlled studies. There are strong arguments to avoid emergency use of agents until trials are completed and analyzed, but the agents suggested are not new. Most are drugs like ivermectin, colchicine, and the interferons, marketed and available for other conditions and with well-known safety profiles. There is a clear need to offer outpatient therapeutic intervention now to the World’s population.

There is a precedent to resolve the conflict between immediate clinical need and the requirements for rigorous controlled trials to prove efficacy. We should remember that the greatest success in fighting a pandemic occurred over the past two decades in the battle against the HIV virus which causes AIDS. AIDS was first recognized in 1981 in the MSM (men who have sex with men) community. The disease was considered a death sentence.

There was widespread fear because there was no treatment, and projections of infection escalated into the millions. The first AIDS remedy was azidothymidine (AZT), synthesized in 1964 in the hope that it would combat cancer. Twenty years later Dr. Samuel Broder, head of the National Cancer Institute, showed that the drug had activity against the HIV virus in vitro (113). Burroughs Wellcome launched a rapidly conceived trial with just 300 patients. They stopped the trial in 16 weeks claiming that more patients survived on AZT. The FDA came under enormous pressure from AIDS activists to make the drug available, and it was approved on March 19, 1987, with only that one trial. It had taken 20 months for the FDA to give approval to release the drug. To this day, the design and results of the trial remain controversial.

The LGBT community continued to battle for early release of other medications to combat the AIDS pandemic. On October 11, 1988, a massive protest occurred at the FDA. It was back then Dr. Anthony Fauci who publicly advanced the idea of a parallel track to make drugs widely available even while studies are progressing: “*Clearly, the standard approach to the design of clinical trials—that is, rigid eligibility criteria as well as the strict regulatory aspects that attend clinical trial investigations and drug approval—was not well-suited to a novel, largely fatal disease such as this with no effective treatments, and we had many intense discussions about how to make that approach more flexible and ethically sound*. *One example, which I and others worked closely with the AIDS activists to develop, was called a parallel track for clinical trials. The parallel track concept, which the United States Food and Drug Administration ultimately came to support, meant that there would be the standard type of highly controlled admission criteria and data collection for the clinical trial of a particular drug. In parallel, however, the drug also could be made available to those who did not meet the trial’s strict admission criteria but were still in dire need of any potentially effective intervention, however unproven, for this deadly disease*” (114).

The parallel track advocated by Dr. Fauci was adopted. Today, there are 41 drugs or combinations approved by the FDA to treat and to prevent HIV infection. There is still no vaccine. There are now an estimated 1.1 million patients with HIV in the United States, most enjoying near normal life expectancy thanks to the antiviral agents. The CDC has contributed greatly to limit the spread of HIV by advocating safe sex practices, but social distancing is not the norm for HIV. Rather “treatment as prevention” for people with HIV using highly active antiretroviral regimens to prevent transmission as well as pre-exposure prophylaxis with a daily antiviral combination pill are currently endorsed by the CDC and adopted in wide segments of the at risk population (115).

In this pandemic crisis, we appeal to public health authorities to change the dynamic of current studies to outpatient care and to cross institutional, commercial and international boundaries to collate and combine all randomized controlled data submitted for all agents in Europe, China, Russia, Japan, India and other countries, and by competing companies, whether officially published, posted on line, or unpublished to finalize confirmatory results. The Solidarity Trial is a model of what could and should be done to unify a worldwide effort to pursue randomized controlled studies in outpatients. At the same time, agents with favorable preliminary results and no safety issues should be made available through a parallel track. In Russia and India, the parallel track has been fully implemented, with FVP now offered as treatment throughout both countries, even as further confirmatory controlled trials proceed. Interferon formulations are now approved treatments in China and Cuba. It is reasonable to authorize vaccination in very large parallel track studies in at risk populations, such as healthcare workers and the elderly. At this time, with no other option available, vaccination will have to be universal, despite the need to perform controlled studies to develop improved vaccines. However, it is unwise to rely solely on the hope for eventual mass vaccination to stop SARS-Cov2. Antiviral medication has succeeded in limiting HIV and hepatitis, and antivirals are just as important as annual vaccination for control of influenza. It is necessary for public health authorities to make hard decisions now despite limited current data and offer outpatient treatments on a broad front with no further delay.

## Data Availability

We have provided a comprehensive list of all clinical trials with their registration as Supplementary Table 1 and all completed trials as Supplementary Table 2

https://www.covid-trials.org/

https://clinicaltrials.gov/

https://www.who.int/ictrp/en/

https://reaganudall.org/covid-19

---

| TRIAL REGISTRATION | COUNTRY | PROJECTED<br>DATE<br>CLOSE ENROLL-MENT | SUBJECTS | AGENTS STUDIED | NUMBER<br>SUBJECTS |
| --- | --- | --- | --- | --- | --- |
| CHICTR2000030946 | China | 2020-Apr | HOSP | Anticoagulants, Other | 120 |
| CHICTR2000029898 | China | 2020-Apr | HOSP | HCQ | 100 |
| NCT04282902 | China | 2020-Apr | HOSP | pirfenidone | 294 |
| CHICTR2000030179 | China | 2020-Apr | HOSP | Plasma based therapy, SOC | 100 |
| CHICTR2000030545 | China | 2020-Apr | HOSP | SOC, Herbal tea | 300 |
| NCT04480593 | Brazil | 2020-Aug | HOSP | Alternative therapy, SOC | 120 |
| NCT04335552 | United States | 2020-Aug | HOSP | AZM, HCQ, SOC | 500 |
| NCT04355637 | Spain | 2020-Aug | HOSP | Corticosteroids, SOC | 300 |
| NCT04349592 | Qatar | 2020-Aug | HOSP | HCQ, HCQ + AZM, SOC | 456 |
| NCT04347031 | Russia | 2020-Aug | HOSP | HCQ, Other, TCZ +AZM | 320 |
| NCT04394442 | Saudi Arabia | 2020-Aug | HOSP | HCQ, SOC | 200 |
| NCT04420247 | Brazil | 2020-Aug | HOSP | HCQ, SOC | 100 |
| NCT04476888 | Pakistan | 2020-Aug | HOSP | Plasma based therapy, SOC | 100 |
| NCT04349098 | United States | 2020-Aug | HOSP | selinexor, SOC | 230 |
| NCT04326426 | United States | 2020-Aug | HOSP | Tradipitant (neurokinin 1 inhibitor),SOC | 300 |
| NCT04445272 | Spain | 2020-Aug | HOSP | TCZ | 500 |
| NCT04311697 | United States | 2020-Aug | HOSP | aviptadil Inhaled solution, SOC | 144 |
| NCT04381858 | Mexico | 2020-Aug | HOSP | Igs, Plasma based therapy | 500 |
| NCT04325906 | United States | 2020-Aug | HOSP | Non-invasive respiratory support, Positioning + Non-invasive res-<br>piratory support | 346 |
| NCT04394377 | Brazil | 2020-Dec | HOSP | Anticoagulants | 600 |
| NCT04351724 | Austria | 2020-Dec | HOSP | Anticoagulants, HCQ, LPV/r, mAb, RAS blockade, SOC | 500 |
| NCT04452565 | United States | 2020-Dec | HOSP | Antiretroviral + Dexamethasone, Other, Other + Antiretroviral,<br>Other + Dexamethasone | 525 |
| NCT04396106 | United States | 2020-Dec | HOSP | AT-527 RNA polymerase inhbitor | 190 |
| NCT04365699 | United States | 2020-Dec | HOSP | AT001, SOC | 500 |
| NCT04321096 | Denmark | 2020-Dec | HOSP | Camostat, SOC | 580 |
| NCT04392531 | Spain | 2020-Dec | HOSP | Cyclosporine, SOC | 120 |
| NCT04397718 | United States | 2020-Dec | HOSP | Hormone therapy, SOC | 198 |
| NCT04421027 | United States | 2020-Dec | HOSP | JAKi, SOC | 600 |
| NCT04469114 | Israel | 2020-Dec | HOSP | JAKi, SOC | 260 |
| NCT04347239 | United States | 2020-Dec | HOSP | Leronlimab, SOC | 390 |
| NCT04376684 | United States | 2020-Dec | HOSP | mAb, SOC | 800 |
| NCT04401202 | Saudi Arabia | 2020-Dec | HOSP | Nigella sativa, SOC | 200 |
| NCT04432298 | United States | 2020-Dec | HOSP | pamrevlumab (Ab to connective tissue growth factor), SOC | 130 |
| NCT04433546 | United States | 2020-Dec | HOSP | pemziviaptadil, SOC | 210 |
| NCT04392232 | United States | 2020-Dec | HOSP | Plasma based therapy | 100 |
| NCT04400058 | United States | 2020-Dec | HOSP | Plasma based therapy, SOC | 208 |
| NCT04359810 | United States | 2020-Dec | HOSP | Plasma based therapy, SOC | 105 |
| NCT04424797 | United States | 2020-Dec | HOSP | Positioning | 100 |
| NCT04391140 | Spain | 2020-Dec | HOSP | Positioning + Respiratory support, Respiratory support | 248 |
| NCT04402879 | Canada | 2020-Dec | HOSP | Positioning, SOC | 596 |
| NCT04365257 | United States | 2020-Dec | HOSP | prazosin, SOC | 220 |
| NCT04412356 | Sweden | 2020-Dec | HOSP | Procedure | 180 |
| NCT04479358 | United States | 2020-Dec | HOSP | SOC, TCZ | 332 |
| NCT04317092 | Italy | 2020-Dec | HOSP | TCZ | 400 |
| NCT04433910 | Germany | 2020-Dec | HOSP | Plasma based therapy, SOC | 106 |
| NCT04433910 | Germany | 2020-Dec | HOSP | Plasma based therapy, SOC | 106 |
| NCT04406389 | United States | 2020-Dec | HOSP | Anticoagulants | 186 |
| NCT04344730 | France | 2020-Dec | HOSP | Dexamethasone with or without respiratory support, SOC | 550 |
| NCT04480424 | United States | 2020-Dec | HOSP | Ivlg, SOC | 100 |
| NCT04443881 | Spain | 2020-Dec | HOSP | Anakinra, SOC | 180 |
| NCT04485429 | Brazil | 2020-Dec | HOSP | Anticoagulants, Anticoagulants + Corticosteroids, Corticosteroids, SOC | 268 |
| NCT04466670 | Brazil | 2020-Dec | HOSP | Anticoagulants, Antihypertensive | 310 |
| NCT04409509 | United States | 2020-Dec | HOSP | FX12 antibody, SOC | 124 |
| NCT04535063 | Argentina | 2020-Dec | HOSP | Plasma based therapy | 200 |
| NCT04336462 | China | 2020-Feb | HOSP | Hydrogen, oxygen, SOC | 100 |
| CHICTR2000032367 | China | 2020-Jul | HOSP | Exercise, SOC | 186 |
| NCT04355897 | United States | 2020-Jul | HOSP | Plasma based therapy | 100 |
| NCT04342182 | Netherlands | 2020-Jul | HOSP | Plasma based therapy, SOC | 426 |
| NCT04345523 | Spain | 2020-Jul | HOSP | Plasma based therapy, SOC | 278 |
| NCT04468646 | Pakistan | 2020-Jul | HOSP | Neurokinin 1 receptor agonist, SOC | 100 |
| CHICTR2000030333 | China | 2020-Jul | HOSP | pirfenidone, SOC | 292 |
| NCT04459325 | Russia | 2020-Jul | HOSP | dornase, alfa | 100 |
| NCT04322344 | Italy | 2020-Jun | HOSP | Alternative therapy, SOC | 120 |
| NCT04305106 | China | 2020-Jun | HOSP | Bevacizumab, SOC | 140 |
| NCT04375202 | Italy | 2020-Jun | HOSP | Colchicine, SOC | 308 |
| NCT04347980 | France | 2020-Jun | HOSP | Dexamethasone + HCQ, HCQ | 122 |
| CHICTR2000029777 | China | 2020-Jun | HOSP | SOC, Herbal tea | 160 |
| CHICTR2000029781 | China | 2020-Jun | HOSP | SOC, Herbal tea | 160 |
| CHICTR2000034796 | China | 2020-Mar | HOSP | Anticoagulants, SOC | 160 |
| NCT04329195 | France | 2020-May | HOSP | Antihypertensive | 554 |
| NCT04329650 | Spain | 2020-May | HOSP | Corticosteroids, Siltuximab | 200 |
| CHICTR2000030894 | China | 2020-May | HOSP | FPV, FPV + TCZ, TCZ | 150 |
| CHICTR2000029603 | China | 2020-May | HOSP | LPV/r, ASC09/r | 160 |
| CHICTR2000030001 | China | 2020-May | HOSP | LPV/r, Emcetrabine/r, SOC | 240 |
| CHICTR2000029763 | China | 2020-May | HOSP | SOC, Heral Tea | 408 |
| CHICTR2000029755 | China | 2020-May | HOSP | SOC, Herbal tea | 120 |
| CHICTR2000029765 | China | 2020-May | HOSP | SOC, TCZ | 188 |
| CHICTR2000030779 | China | 2020-May | HOSP | ulinastatin, SOC | 100 |
| NCT04310228 | China | 2020-May | HOSP | FPV, FPV + TCZ, TCZ | 150 |
| NCT04346199 | Argentina | 2020-Nov | HOSP | Acalabrutinib, SOC | 140 |
| NCT04444700 | Brazil | 2020-Nov | HOSP | Anticoagulants, SOC | 462 |
| NCT04362085 | Canada | 2020-Nov | HOSP | Anticoagulants, SOC | 462 |
| NCT04344288 | France | 2020-Nov | HOSP | Corticosteroids, SOC | 304 |
| NCT04382053 | Brazil | 2020-Nov | HOSP | DFV890, SOC | 120 |
| NCT04358081 | United States | 2020-Nov | HOSP | HCQ, HCQ + AZM, SOC | 444 |
| NCT04362137 | United States | 2020-Nov | HOSP | JAKi, SOC | 402 |
| NCT04511819 | United States | 2020-Nov | HOSP | Losmapimod, SOC | 410 |
| NCT04448756 | United States | 2020-Nov | HOSP | M5049 (toll like receptor inhibitor), Other | 150 |
| NCT04441385 | Spain | 2020-Nov | HOSP | Maraviroc, SOC | 200 |
| NCT04463420 | Iran | 2020-Nov | HOSP | PHR160 asal spray+ HCQ + NSAIDs + AZM | 224 |
| NCT04388410 | Mexico | 2020-Nov | HOSP | Plasma based therapy, SOC | 410 |
| NCT04495101 | Spain | 2020-Nov | HOSP | prolactin (proteinase inhibitor), SOC | 100 |
| NCT04449718 | Brazil | 2020-Nov | HOSP | SOC, Vitamin D | 200 |
| NCT04386850 | Iran | 2020-Nov | HOSP | SOC, Vitamins | 1500 |
| NCT04384900 | Denmark | 2020-Nov | HOSP | Positioning | 150 |
| NCT04400890 | United States | 2020-Nov | HOSP | Plant polyphenols | 200 |
| NCT04477655 | Mexico | 2020-Nov | HOSP | Positioning, SOC | 200 |
| NCT04427098 | Italy | 2020-Oct | HOSP | Anticoagulants | 300 |
| NCT04530578 | Argentina | 2020-Oct | HOSP | Anticoagulants | 200 |
| NCT04486508 | Iran | 2020-Oct | HOSP | Anticoagulants, SOC, Statins | 600 |
| NCT04470622 | United States | 2020-Oct | HOSP | Aprepitant (antiemetic), SOC | 100 |
| NCT04340557 | United States | 2020-Oct | HOSP | ARB, SOC | 200 |
| NCT04364893 | Brazil | 2020-Oct | HOSP | ARBs + ACEi, SOC | 700 |
| NCT04350320 | Spain | 2020-Oct | HOSP | Colchicine, SOC | 102 |
| NCT04322396 | Denmark | 2020-Oct | HOSP | HCQ + AZM, SOC | 226 |
| NCT04348656 | United States | 2020-Oct | HOSP | Plasma based therapy, SOC | 1200 |
| NCT04385043 | Italy | 2020-Oct | HOSP | Plasma based therapy, SOC | 400 |
| NCT04395144 | Canada | 2020-Oct | HOSP | Positioning, SOC | 346 |
| NCT04335071 | Switzerland | 2020-Oct | HOSP | SOC, TCZ | 100 |
| NCT04385836 | Saudi Arabia | 2020-Sep | HOSP | alpha1 antitrypsin, SOC | 150 |
| NCT04341116 | United States | 2020-Sep | HOSP | anti GM-CSF, SOC | 144 |
| NCT04335136 | Austria | 2020-Sep | HOSP | Antihypertensive, SOC | 200 |
| NCT04386616 | United States | 2020-Sep | HOSP | astegolimab, SOC | 300 |
| NCT04452435 | India | 2020-Sep | HOSP | C21, SOC | 100 |
| NCT04451174 | Chile | 2020-Sep | HOSP | Corticosteroids, SOC | 184 |
| NCT04332094 | Spain | 2020-Sep | HOSP | HCQ + AZM, TCZ + AZM + HCQ | 276 |
| NCT04403100 | Brazil | 2020-Sep | HOSP | HCQ, HCQ + LPV/r, LPV/r, SOC | 1968 |
| NCT04370262 | United States | 2020-Sep | HOSP | HCQ, HCQ + Other, SOC | 942 |
| NCT04351152 | United States | 2020-Sep | HOSP | Lenzilumab, SOC | 300 |
| NCT04334629 | United Kingdom | 2020-Sep | HOSP | NSAIDs, SOC | 230 |
| NCT04374526 | Italy | 2020-Sep | HOSP | Plasma based therapy, SOC | 182 |
| CHICTR2000030522 | China | 2020-Sep | HOSP | SOC, Herbal tea | 100 |
| NCT04379271 | Bulgaria | 2020-Sep | HOSP | vidofludimus (pyrimidine synthesis inhibitor), SC | 230 |
| NCT04377620 | United States | 2020-Sep | HOSP | JAKi, SOC | 500 |
| NCT04445935 | Qatar | 2020-Sep | HOSP | Anticoagulants, SOC | 100 |
| NCT04359277 | United States | 2021-Apr | HOSP | Anticoagulants | 1000 |
| NCT04360824 | United States | 2021-Apr | HOSP | Anticoagulants | 170 |
| NCT04360824 | United States | 2021-Apr | HOSP | Anticoagulants | 170 |
| NCT04312009 | United States | 2021-Apr | HOSP | ARBs | 200 |
| NCT04381377 | Russia | 2021-Apr | HOSP | Azoximer bromide, SOC | 394 |
| NCT04367168 | Mexico | 2021-Apr | HOSP | Colchicine, SOC | 174 |
| IRCT20200517047485N1 | Iran | 2021-Apr | HOSP | HCQ + Atazanavir/ritonavir, HCQ + LPV/r | 108 |
| NCT04328493 | Vietnam | 2021-Apr | HOSP | HCQ, SOC | 250 |
| NCT04328493 | Vietnam | 2021-Apr | HOSP | HCQ, SOC | 250 |
| NCT04320056 | Canada | 2021-Apr | HOSP | Non-invasive respiratory support, SOC | 216 |
| NCT04344431 | France | 2021-Apr | HOSP | Non-invasive respiratory support, SOC | 100 |
| NCT04362176 | United States | 2021-Apr | HOSP | Plasma based therapy, SOC | 500 |
| NCT04359797 | United States | 2021-Apr | HOSP | Positioning | 300 |
| NCT04346615 | United States | 2021-Apr | HOSP | Zavegepant (CGRP agonist), SOC | 120 |
| NCT04352751 | Pakistan | 2021-Apr | HOSP | Plasma based therapy | 2000 |
| IRCT20200414047072N1 | Iran | 2021-Apr | HOSP | Plasma based therapy | 357 |
| NCT04389710 | United States | 2021-Apr | HOSP | Plasma based therapy | 100 |
| NCT04355143 | United States | 2021-Apr | HOSP | Colchicine, SOC | 150 |
| NCT04387760 | Bahrain | 2021-Apr | HOSP | FPV, HCQ, SOC | 150 |
| CHICTR2000029739 | China | 2021-Aug | HOSP | Non-invasive respiratory support | 440 |
| NCT04513158 | United States | 2021-Aug | HOSP | Plasma based therapy | 100 |
| NCT04345692 | United States | 2021-Dec | HOSP | HCQ, SOC | 350 |
| NCT04381936 | United Kingdom | 2021-Dec | HOSP | RECOVERY TRIAL Conv plasma vs TCZ vs Monoclonal Abs | 15000 |
| CHICTR2000031430 | China | 2021-Dec | HOSP | SOC, Stem cells | 200 |
| NCT04412772 | United States | 2021-Dec | HOSP | SOC, TCZ | 300 |
| NCT04339816 | Czechia | 2021-Dec | HOSP | HCQ, | 240 |
| NCT04447469 | United States | 2021-Feb | HOSP | Antibody to GM-CSF | 573 |
| NCT04390594 | Senegal | 2021-Feb | HOSP | HCQ, HCQ + AZM | 186 |
| NCT04477993 | Brazil | 2021-Feb | HOSP | JAKi, SOC | 200 |
| NCT04393246 | United Kingdom | 2021-Feb | HOSP | Other, SGLT2 + Other, SOC | 1407 |
| CHICTR2000029757 | China | 2021-Feb | HOSP | Plasma based therapy, SOC | 200 |
| NCT04348383 | Spain | 2021-Jan | HOSP | Antithrombotics, SOC | 120 |
| NCT04338958 | Germany | 2021-Jan | HOSP | JAKi | 200 |
| NCT04426695 | United States | 2021-Jan | HOSP | mAB | 2970 |
| NCT04460183 | United Kingdom | 2021-Jan | HOSP | Non-invasive respiratory support, SOC | 300 |
| CHICTR2000029806 | China | 2021-Jan | HOSP | PD1, thymosin, SOC | 120 |
| NCT04439071 | United States | 2021-Jan | HOSP | PTC299 (DDODH inhibitor), SOC | 380 |
| NCT04363814 | Dominican Re-public | 2021-Jul | HOSP | BAKTEC-R bacterial identification, SOC | 100 |
| NCT04514627 | United States | 2021-Jul | HOSP | Other, SOC | 100 |
| NCT04401423 | United States | 2021-Jun | HOSP | Antihypertensive, SOC | 100 |
| NCT04359953 | France | 2021-Jun | HOSP | ARBs, AZM, HCQ | 1600 |
| NCT04414631 | Switzerland | 2021-Jun | HOSP | Conestat alfa (C1esterase inhibitor), SOC | 120 |
| NCT04345289 | Denmark | 2021-Jun | HOSP | Conv Plasma | 1100 |
| NCT04432272 | United States | 2021-Jun | HOSP | Plasma based therapy | 500 |
| NCT04418518 | United States | 2021-Jun | HOSP | Plasma-based therapy, SOC | 1200 |
| NCT04364763 | United States | 2021-Jun | HOSP | RBT-9 (antiviral), SOC | 252 |
| NCT04322773 | Denmark | 2021-Jun | HOSP | Sarilumab, SOC, TCZ | 200 |
| NCT04323514 | Italy | 2021-Mar | HOSP | Axcorbic Acid | 500 |
| IRCT20170731035423N2 | Iran | 2021-Mar | HOSP | BCC1 and Hep S, SOC | 120 |
| NCT04420299 | Spain | 2021-Mar | HOSP | Bemiparin (low molecular wt heparin), SOC | 120 |
| NCT04351763 | Poland | 2021-Mar | HOSP | CCB, Other, SOC | 804 |
| NCT04476979 | France | 2021-Mar | HOSP | Corticosteroids, TCZ + Corticosteroids | 120 |
| NCT04373733 | United Kingdom | 2021-Mar | HOSP | FPV, HCQ + AZM + Zinc, SOC | 450 |
| NCT04342221 | Germany | 2021-Mar | HOSP | HCQ, SOC | 220 |
| IRCT20170922036314N4 | Iran | 2021-Mar | HOSP | Psychological therapy, SOC | 112 |
| NCT04380961 | United States | 2021-Mar | HOSP | sirukumab (IL6 inhibitor), SOC | 270 |
| NCT04306393 | United States | 2021-Mar | HOSP | Non-invasive respiratory support, SOC | 200 |
| NCT04306393 | United States | 2021-Mar | HOSP | Non-invasive respiratory support, SOC | 200 |
| NCT04306393 | United States | 2021-Mar | HOSP | Non-invasive respiratory support, SOC | 200 |
| NCT04376034 | United States | 2021-Mar | HOSP | Plasma based therapy, SOC | 240 |
| NCT04366050 | United States | 2021-May | HOSP | ACEi, SOC | 560 |
| NCT04379492 | United States | 2021-May | HOSP | HCQ, SOC | 120 |
| NCT04333420 | Netherlands | 2021-May | HOSP | IFN, SOC | 390 |
| NCT04390464 | United Kingdom | 2021-May | HOSP | JAKi, mAb, SOC | 1167 |
| NCT04365985 | United States | 2021-May | HOSP | Opiate antagonists, Opiate antagonists + Anesthetic, SOC | 500 |
| NCT04516915 | United Kingdom | 2021-May | HOSP | Oseltamivir, vidoflodimus + Oseltamivir | 120 |
| IRCT20200501047258N1 | Iran | 2021-May | HOSP | Plasma based therapy, SOC | 120 |
| NCT04374539 | Spain | 2021-May | HOSP | Plasma based therapy, SOC | 116 |
| NCT04367077 | United States | 2021-Sep | HOSP | SOC, Stem cells | 400 |
| NCT04415086 | Brazil | 2022-Apr | HOSP | Plasma-based therapy, SOC | 120 |
| NCT04478071 | United States | 2022-Aug | HOSP | Vadadustat(hypoxia inducible factor), SOC | 300 |
| NCT04321993 | Canada | 2022-Feb | HOSP | HCQ, JAKi, LPV/r, Sarilumab | 800 |
| NCT04394416 | United States | 2022-Jun | HOSP | Imatinib, SOC | 204 |
| NCT04330690 | Canada | 2022-Mar | HOSP | LPV/r, SOC | 2900 |
| NCT04354831 | United States | 2022-May | HOSP | Plasma based therapy | 131 |
| NCT04412486 | United States | 2022-May | HOSP | Plasma based therapy | 100 |
| NCT04470544 | United States | 2022-Sep | HOSP | Camostat, SOC | 264 |
| NCT04364737 | United States | 2023-Jan | HOSP | Plasma based therapy, SOC | 300 |
| NCT04276896 | China | 2023-Jul | HOSP | LV-SMENP-DC Vaccine | 100 |
| NCT04415060 | Canada | 2023-Jun | HOSP | Anesthetic, SOC | 752 |
| NCT04315948 | Austria | 2023-Mar | HOSP | HCQ, LPV/r, LPV/r + IFN, Remdesivir, SOC | 3100 |
| NCT04435353 | Kazakhstan | NA | HOSP | Alternative therapy | 200 |
| CTRI/2020/05/025434 | India | NA | HOSP | Alternative therapy | 135 |
| CTRI/2020/06/025977 | India | NA | HOSP | Alternative therapy, SOC | 120 |
| CTRI/2020/06/025590 | India | NA | HOSP | Alternative therapy, SOC | 120 |
| CTRI/2020/04/024883 | India | NA | HOSP | Alternative therapy, SOC | 112 |
| NCT04347226 | United States | NA | HOSP | anti-IL8, SOC | 138 |
| NCT04401293 | United States | NA | HOSP | Anticoagulants | 308 |
| 2020-001972-13 | Italy | NA | HOSP | Anticoagulants | 300 |
| NCT04508439 | Mexico | NA | HOSP | Anticoagulants | 130 |
| NCT04372589 | United States | NA | HOSP | Anticoagulants, SOC | 3000 |
| NCT04389840 | United States | NA | HOSP | Anticoagulants, SOC | 524 |
| ACTRN12620000517976 | Australia | NA | HOSP | Anticoagulants, SOC | 206 |
| NCT04493359 | Brazil | NA | HOSP | Antihypertensive | 240 |
| NCT04328012 | United States | NA | HOSP | ARBs, HCQ, LPV/r, SOC | 4000 |
| ISRCTN48734830 | Sweden | NA | HOSP | ARBs, SOC | 750 |
| NCT04335786 | Netherlands | NA | HOSP | ARBs, SOC | 651 |
| CTRI/2020/05/025319 | India | NA | HOSP | ARBs, SOC | 186 |
| CTRI/2020/05/025271 | India | NA | HOSP | BCG, SOC | 480 |
| NCT04334460 | United States | NA | HOSP | BLD2660 (protease inhibitor), SOC | 120 |
| CTRI/2020/06/026192 | India | NA | HOSP | C21, SOC | 100 |
| NCT04362813 | United States | NA | HOSP | Canakinumab, SOC | 450 |
| NCT04328480 | Argentina | NA | HOSP | Colchicine, SOC | 2500 |
| NCT04381364 | Sweden | NA | HOSP | Corticosteroids, SOC | 446 |
| NCT04331054 | France | NA | HOSP | Corticosteroids, SOC | 436 |
| NCT04323592 | Italy | NA | HOSP | Corticosteroids, SOC | 104 |
| NCT04345614 | United States | NA | HOSP | CRAC channel inhibitor, SOC | 400 |
| NCT04395105 | Argentina | NA | HOSP | Dexamethasone, SOC | 284 |
| NCT04424901 | United States | NA | HOSP | Dipyridamole, SOC | 100 |
| 2020-001643-13 | United Kingdom | NA | HOSP | DPP1 inhibitor, SOC | 300 |
| ISRCTN30564012 | United Kingdom | NA | HOSP | DPP1 inhibitor, SOC | 300 |
| NCT04494984 | Argentina | NA | HOSP | Equine hyperimmune serum, SOC | 242 |
| JPRN-JRCTS031190226 | Japan | NA | HOSP | FPV | 100 |
| NCT04411433 | Turkey | NA | HOSP | FPV, FPV + AZM, HCQ, HCQ + AZM, HCQ + FPV | 1000 |
| CTRI/2020/06/025957 | India | NA | HOSP | FPV, FPV + Umifenovir | 158 |
| 2020-001904-41 | United Kingdom | NA | HOSP | FPV, SOC | 302 |
| CTRI/2020/06/025704 | India | NA | HOSP | HCQ + AZM, HCQ + Other + AZM | 120 |
| NCT04355052 | Israel | NA | HOSP | HCQ + AZM, HCQ + Other, SOC | 250 |
| NCT04392973 | Saudi Arabia | NA | HOSP | HCQ + FPV, SOC | 520 |
| NCT04346147 | Spain | NA | HOSP | HCQ + Imatinib, HCQ + JAKi, HCQ + LPV/r | 165 |
| 2020-002035-30 | Italy | NA | HOSP | HCQ, HCQ + JAKi | 116 |
| 2020-000936-23 | Austria | NA | HOSP | HCQ, LPV/r, Remdesivir | 3100 |
| PACTR202004801273802 | Nigeria | NA | HOSP | HCQ, SOC | 800 |
| ISRCTN88057279 | United Kingdom | NA | HOSP | HCQ, SOC | 500 |
| NCT04459676 | Brazil | NA | HOSP | heptocyte growth factor, SOC | 100 |
| NCT04382924 | United States | NA | HOSP | Ifenprodil, SOC | 682 |
| 2020-001023-14 | United Kingdom | NA | HOSP | IFN, SOC | 400 |
| NL8491 | Netherlands | NA | HOSP | imatinib, SOC | 386 |
| CTRI/2020/04/024806 | India | NA | HOSP | Imatinib, SOC | 100 |
| 2020-001750-22 | United Kingdom | NA | HOSP | Immunomodulators, JAKi, SOC | 186 |
| NCT04402866 | United Kingdom | NA | HOSP | JAK1,SOC | 159 |
| NCT04373044 | United States | NA | HOSP | JAKi, HCQ, SOC | 144 |
| ISRCTN11188345 | United Kingdom | NA | HOSP | JAKi, mAb, SOC | 375 |
| NCT04404361 | United States | NA | HOSP | JAKi, SOC | 358 |
| 2020-001807-18 | Finland | NA | HOSP | JAKi, SOC | 159 |
| 2020-001759-42 | Argentina | NA | HOSP | mAb, SOC | 800 |
| NCT04382651 | United States | NA | HOSP | MAS825, SOC | 120 |
| NCT04374032 | Bosnia & Herzegovina | NA | HOSP | metenkefalin + tridecactide,SOC | 120 |
| ISRCTN40580903 | United Kingdom | NA | HOSP | Mylotarg, Infliximab, Namilumab, SOC | 588 |
| 2020-001236-10 | Netherlands | NA | HOSP | NA | 304 |
| NCT04395807 | Sweden | NA | HOSP | Non-invasive respiratory support | 120 |
| NCT04467840 | Israel | NA | HOSP | opaganib (sphingosine kinase), SOC | 270 |
| 2020-001805-21 | United Kingdom | NA | HOSP | Oseltamivir, Oseltamivir + Other | 120 |
| 2020-002229-27 | United Kingdom | NA | HOSP | Other, Other + SGLT2, SOC | 1407 |
| ISRCTN60069084 | Oman | NA | HOSP | Other, SOC | 1180 |
| NCT04472728 | United States | NA | HOSP | Other, SOC | 310 |
| NCT04408040 | United States | NA | HOSP | Plasma based therapy | 700 |
| NCT04393727 | Italy | NA | HOSP | Plasma based therapy | 126 |
| NCT04344535 | United States | NA | HOSP | Plasma based therapy, SOC | 500 |
| NCT04429854 | Belgium | NA | HOSP | Plasma based therapy, SOC | 483 |
| NL8633 | Netherlands | NA | HOSP | Plasma based therapy, SOC | 430 |
| NCT04479163 | Argentina | NA | HOSP | Plasma based therapy, SOC | 210 |
| ISRCTN85216856 | Ecuador | NA | HOSP | Plasma based therapy, SOC | 200 |
| NCT04358939 | France | NA | HOSP | Positioning | 248 |
| NCT04366856 | France | NA | HOSP | Positioning, SOC | 500 |
| NCT04383613 | Canada | NA | HOSP | Positioning, SOC | 350 |
| ISRCTN54917435 | Sweden | NA | HOSP | Positioning, SOC | 240 |
| NCT04343963 | Mexico | NA | HOSP | Pyridostigmine, SOC | 436 |
| 2020-001497-30 | France | NA | HOSP | ravilizumab(complement inhibitor), SOC | 270 |
| 2020-001786-36 | Belgium | NA | HOSP | Recombinant interleukin 7, SOC | 200 |
| 2020-001573-78 | United States | NA | HOSP | Recombinant interleukin 7, SOC | 200 |
| 2020-001453-49 | France | NA | HOSP | Remdesivir | 2500 |
| NCT04409262 | United States | NA | HOSP | Remdesivir, Remdesivir + TCZ | 450 |
| 2020-001052-18 | Denmark | NA | HOSP | Remdesivir, SOC | 440 |
| ISRCTN13035264 | Denmark | NA | HOSP | Remdesivir, SOC | 400 |
| NCT04359901 | United States | NA | HOSP | Sarilumab, SOC | 120 |
| 2020-001473-79 | Italy | NA | HOSP | SGLT2, SOC | 900 |
| NCT04389450 | United States | NA | HOSP | SOC, Stem cells | 140 |
| CHICTR2000029990 | China | NA | HOSP | SOC, Stem cells | 120 |
| NCT04366271 | Spain | NA | HOSP | SOC, Stem cells | 106 |
| NCT04393415 | Egypt | NA | HOSP | SOC, Stem cells | 100 |
| CTRI/2020/05/025369 | India | NA | HOSP | SOC, TCZ | 180 |
| NCT04411446 | Argentina | NA | HOSP | SOC, Vitamins | 1265 |
| ACTRN12620000454976 | Australia | NA | HOSP | SOC, Zinc | 160 |
| 2020-002120-37 | France | NA | HOSP | sodium nitrite, SOC | 300 |
| 2020-002060-31 | Czechia | NA | HOSP | SOLIDARITY TRIAL | 100000 |
| ISRCTN83971151 | Argentina | NA | HOSP | SOLIDARITY TRIAL | 10000 |
| CTRI/2020/04/024773 | Argentina | NA | HOSP | SOLIDARITY TRIAL | 7000 |
| NCT04417257 | United States | NA | HOSP | tenoretidine, SOC | 240 |
| NCT04419623 | United States | NA | HOSP | TL-895 (tyrosine kinase inhibitor), SOC | 146 |
| NL8504 | Netherlands | NA | HOSP | TCZ | 354 |
| NCT04378920 | France | NA | HOSP | trancrocetin (NMDA inhibitor), SOC | 180 |
| 2020-001264-28 | Bulgaria | NA | HOSP | vidofludimus (pyrimidine synthesis inhibitor), SC | 200 |
| NCT04453384 | France | NA | HOSP | XAV19( spike protein antibody), SOC | 368 |
| NCT04373707 | France | NA | HOSP | Anticoagulants | 602 |
| NCT04345848 | Switzerland | NA | HOSP | Anticoagulants | 200 |
| CTRI/2020/06/026193 | India | NA | HOSP | HCQ +/- AZM, HCQ + Lithium +/- AZM | 100 |
| NCT04321616 | Norway | NA | HOSP | HCQ, Remdesivir, SOC | 700 |
| 2020-001754-21 | France | NA | HOSP | JAKi + TCZ or Anakinra, SOC, TCZ or Anakinra | 150 |
| NCT04346797 | France | NA | HOSP | mAb, SOC | 120 |
| NCT04347681 | Saudi Arabia | NA | HOSP | Plasma based therapy | 575 |
| NCT04474340 | Kuwait | NA | HOSP | Plasma based therapy, SOC | 300 |
| NCT04361253 | United States | NA | HOSP | Plasma based therapy, SOC | 220 |
| NCT04401150 | Canada | NA | HOSP | SOC, Vitamins | 800 |
| NCT04498936 | Egypt | NA | HOSP | Sofosbuvir/Ledipasvir and Nitazoxanide, DAA, SOC | 240 |
| NCT04363853 | Mexico | NA | HOSP | TCZ | 200 |
| 2020-001644-25 | Argentina | NA | HOSP | Acalabrutinib, SOC | 177 |
| NCT04367831 | United States | NA | HOSP | Anticoagulants | 100 |
| NCT04409834 | United States | NA | HOSP | Anticoagulants, Anticoagulants + Antiplatelets | 750 |
| 2020-001921-30 | Italy | NA | HOSP | Anticoagulants, Corticosteroids + Anticoagulants | 200 |
| NCT04371367 | France | NA | HOSP | Avdirolimab (anti C5 antibody), SOC | 108 |
| NCT04355364 | France | NA | HOSP | dornase alfa, SOC | 100 |
| NCT04432051 | Turkey | NA | HOSP | Monitoring, SOC | 100 |
| NCT04358003 | United States | NA | HOSP | Plasma based therapy | 2000 |
| NCT04350580 | France | NA | HOSP | Polyvalent Immunoglobulin, SOC | 138 |
| NCT04347941 | Ireland | NA | HOSP | Positioning, SOC | 200 |
| JPRN-JRCT2031200126 | United States | NA | HOSP | ravilzumab (complement inhibitor), SOC | 270 |
| NCT04371393 | United States | NA | HOSP | SOC, Stem cells | 300 |
| DRKS00021238 | Germany | NA | HOSP | SOC, TCZ | 200 |
| NCT04264533 | China | NA | HOSP | SOC, Vitamin C | 140 |
| NCT04364009 | France | NA | HOSP | Anakinra, SOC | 240 |
| JPRN-JRCTS031200026 | Japan | NA | HOSP | Anticoagulants, FPV | 160 |
| PACTR202007606032743 | Egypt | NA | HOSP | Anticoagulants, SOC | 100 |
| NCT04263402 | China | NA | HOSP | Corticosteroids | 100 |
| KCT0005105 | South Korea | NA | HOSP | Corticosteroids, HCQ + Corticosteroids, SOC | 141 |
| NCT04361474 | France | NA | HOSP | Corticosteroids, SOC | 120 |
| NCT04530409 | Egypt | NA | HOSP | Dexamethasone | 450 |
| NCT04384588 | Chile | NA | HOSP | Plasma based therapy | 100 |
| CTRI/2020/06/025803 | India | NA | HOSP | Plasma based therapy, SOC | 400 |
| NCT04425915 | India | NA | HOSP | Plasma based therapy, SOC | 400 |
| NCT04345419 | Egypt | NA | HOSP | remdesivir, chloroquine, SOC | 120 |
| 2020-001390-76 | Italy | NA | HOSP | Sarilumab, SOC | 171 |
| NCT04486144 | United States | NA | OUTPAT | Alternative therapy, SOC | 100 |
| CHICTR2000029899 | China | 2020-Apr | OUTPAT | HCQ | 100 |
| NCT04438850 | Italy | 2020-Aug | OUTPAT | Ivermectin, SOC | 102 |
| NCT04337918 | Canada | 2020-Aug | OUTPAT | Non-invasive respiratory support | 200 |
| NCT04369365 | Austria | 2020-Aug | OUTPAT | AZM, SOC | 200 |
| CHICTR2000029601 | China | 2020-Aug | OUTPAT | Education, Education + Herbal tea | 400 |
| NCT04466540 | Brazil | 2020-Aug | OUTPAT | HCQ, SOC | 1300 |
| NCT04466540 | Brazil | 2020-Aug | OUTPAT | HCQ, SOC | 1300 |
| NCT04438057 | United States | 2020-Aug | OUTPAT | Plasma based therapy, SOC | 150 |
| NCT04406246 | Mexico | 2020-Dec | OUTPAT | Niazoxanide, SOC | 150 |
| NCT04356495 | France | 2020-Dec | OUTPAT | Antihypertensive, FPV, HCQ, Other, SOC | 634 |
| NCT04532931 | South Africa | 2020-Dec | OUTPAT | Antimalarial, DAA, FPV + Antivirals, SOC | 250 |
| NCT04322682 | United States | 2020-Dec | OUTPAT | Colchicine, SOC | 6000 |
| NCT04322565 | Italy | 2020-Dec | OUTPAT | Colchicine, SOC | 310 |
| NCT04334382 | United States | 2020-Dec | OUTPAT | HCQ vs AZM | 1550 |
| CHICTR2000029741 | China | 2020-Dec | OUTPAT | HCQ, LPV/r | 112 |
| NCT04372628 | United States | 2020-Dec | OUTPAT | HCQ, LPV/r, SOC | 600 |
| NCT04529525 | Argentina | 2020-Dec | OUTPAT | Ivermectin, SOC | 500 |
| NCT04446377 | United States | 2020-Dec | OUTPAT | LAM002 (lysosome activator), SOC | 142 |
| NCT04425629 | United States | 2020-Dec | OUTPAT | mAB, SOC | 2104 |
| NCT04342728 | United States | 2020-Dec | OUTPAT | SOC, Ascorbic Acid, Ascorbic Acid + Zinc, Zinc | 520 |
| CHICTR2000029621 | China | 2020-Dec | OUTPAT | SOC, Umifenovir | 380 |
| NCT04338074 | United States | 2020-Dec | OUTPAT | tranexamic acid | 100 |
| NCT04464408 | Saudi Arabia | 2020-Dec | OUTPAT | FPV, SOC | 576 |
| CHICTR2000029780 | China | 2020-Jun | OUTPAT | SOC, Herbal tea | 160 |
| NCT04346667 | Pakistan | 2020-May | OUTPAT | HCQ, SOC | 400 |
| NCT04351191 | Pakistan | 2020-May | OUTPAT | HCQ, SOC | 400 |
| NCT04345991 | France | 2020-May | OUTPAT | Plasma based therapy, SOC | 120 |
| NCT04416399 | United Kingdom | 2020-Nov | OUTPAT | Corticosteroids, SOC | 478 |
| NCT04517422 | Mexico | 2020-Nov | OUTPAT | Nutrition, SOC | 300 |
| NCT04486313 | United States | 2020-Oct | OUTPAT | Antivirals + Vitamins, Vitamins | 800 |
| NCT04363827 | Italy | 2020-Sep | OUTPAT | HCQ, SOC | 2300 |
| NCT04332107 | United States | 2020-Sep | OUTPAT | AZM, SOC | 2271 |
| NCT04381962 | United Kingdom | 2020-Sep | OUTPAT | AZM, SOC | 800 |
| NCT04416334 | Spain | 2020-Sep | OUTPAT | Colchicine, SOC | 1028 |
| NCT04377711 | United States | 2020-Sep | OUTPAT | Corticosteroids, SOC | 400 |
| NCT04370782 | United States | 2020-Sep | OUTPAT | HCQ + AZM+ Zinc, HCQ + Zinc + Antibiotics | 750 |
| NCT04331600 | Poland | 2020-Sep | OUTPAT | HCQ, SOC | 400 |
| NCT04518410 | United States | 2020-Sep | OUTPAT | Other, SOC | 2000 |
| CHICTR2000033057 | China | 2020-Sep | OUTPAT | Rehabilitation | 160 |
| NCT04427501 | United States | 2020-Sep | OUTPAT | mAB, SOC | 800 |
| NCT04329923 | United States | 2021-Apr | OUTPAT | HCQ, SOC | 400 |
| NCT04390503 | United States | 2021-Apr | OUTPAT | Plasma based therapy, SOC | 200 |
| NCT04311177 | United States | 2021-Apr | OUTPAT | ARBs, SOC | 580 |
| NCT04523090 | South Africa | 2021-Apr | OUTPAT | Nitoxanide | 960 |
| CHICTR2000029589 | China | 2021-Dec | OUTPAT | SOC, Herbal tea | 120 |
| CHICTR2000029539 | China | 2021-Feb | OUTPAT | LPV/r, SOC | 328 |
| NCT04460690 | United States | 2021-Jul | OUTPAT | Rapid SARS-Cov-2 detection | 5000 |
| NCT04475601 | Sweden | 2021-Jul | OUTPAT | Hormone therapy, SOC | 500 |
| NCT04342689 | United States | 2021-Jun | OUTPAT | Nutrition, SOC | 1500 |
| NCT04418505 | United States | 2021-Mar | OUTPAT | Alternative therapy, SOC | 280 |
| NCT04357990 | Iceland | 2021-Mar | OUTPAT | Alternative therapy, SOC | 128 |
| NCT04400799 | Switzerland | 2021-Mar | OUTPAT | Anticoagulants, SOC | 1000 |
| CHICTR2000030804 | China | 2021-Mar | OUTPAT | SOC, Herbal tea | 128 |
| NCT04353284 | United States | 2021-May | OUTPAT | Camostat, SOC | 114 |
| NCT04331899 | United States | 2021-May | OUTPAT | IFN, SOC | 120 |
| NCT04342169 | United States | 2022-Apr | OUTPAT | HCQ, SOC | 400 |
| NCT04373460 | United States | 2022-Dec | OUTPAT | Plasma based therapy, SOC | 1344 |
| NCT04355767 | United States | 2022-Dec | OUTPAT | Plasma based therapy, SOC | 600 |
| CHICTR2000030084 | China | 2022-Feb | OUTPAT | Psychological therapy, SOC | 180 |
| CTRI/2020/05/025490 | India | NA | OUTPAT | Alternative therapy, SOC | 110 |
| NCT04404218 | Canada | NA | OUTPAT | Acai Berry, SOC | 480 |
| ISRCTN13311119 | India | NA | OUTPAT | Alternative therapy, SOC | 116 |
| JPRN-UMIN000040602 | Brazil | NA | OUTPAT | Alternative therapy, SOC | 100 |
| ISRCTN59048638 | Mexico | NA | OUTPAT | Anticoagulants, SOC | 200 |
| NCT04365582 | France | NA | OUTPAT | AZM, HCQ, LPV/r, SOC | 640 |
| NCT04359329 | United States | NA | OUTPAT | Estrogen steroid hormone, SOC | 110 |
| 2020-002106-68 | United Kingdom | NA | OUTPAT | FPV, LPV/r, LPV/r + FPV, SOC | 240 |
| NCT04346628 | United States | NA | OUTPAT | FPV, SOC | 120 |
| ISRCTN86534580 | United Kingdom | NA | OUTPAT | HCQ, SOC | 3000 |
| 2020-001469-35 | Poland | NA | OUTPAT | HCQ, SOC | 400 |
| NCT04351516 | Germany | NA | OUTPAT | HCQ, SOC | 350 |
| 2020-001558-23 | Italy | NA | OUTPAT | HCQ, SOC | 216 |
| NCT04446429 | Brazil | NA | OUTPAT | Hormone therapy + Ivermectin + AZM, Ivermectin + AZM | 381 |
| NCT04382131 | United Kingdom | NA | OUTPAT | Hygiene, SOC | 405 |
| NCT04412018 | Canada | NA | OUTPAT | Icosapent ethyl, SOC | 100 |
| NCT04445311 | Egypt | NA | OUTPAT | Ivermectin, SOC | 100 |
| NCT04414124 | United States | NA | OUTPAT | KB109 (glycan), SOC | 400 |
| NCT04405570 | United States | NA | OUTPAT | molnupiravir )N4-hydroxycytidine derivative), SOC | 108 |
| CHICTR2000029387 | China | NA | OUTPAT | Ribavirin+ IFN, Ribavirin + LPV/r + IFN, LPV/r + IFN | 108 |
| CHICTR2000029855 | China | NA | OUTPAT | SOC, Herbal tea | 180 |
| NCT04482673 | United States | NA | OUTPAT | SOC, Vitamin D | 140 |
| CTRI/2020/05/025167 | India | NA | OUTPAT | Thymoquinone, SOC | 100 |
| CHICTR2000030481 | China | 2020-Apr | OUTPAT,<br>HOSP | Corticosteroids, SOC | 200 |
| CHICTR2000030471 | China | 2020-Apr | OUTPAT,<br>HOSP | Nutrition, SOC | 394 |
| CHICTR2000029518 | China | 2020-Apr | OUTPAT,<br>HOSP | SOC, Herbal tea | 140 |
| CHICTR2000029974 | China | 2020-Aug | OUTPAT,<br>HOSP | SOC, Herbal tea | 300 |
| NCT04467918 | Brazil | 2020-Aug | OUTPAT,<br>HOSP | Alternative therapy, SOC | 100 |
| CHICTR2000033318 | China | 2020-Dec | OUTPAT,<br>HOSP | Exercise, SOC | 104 |
| NCT04529499 | Kuwait | 2020-Dec | OUTPAT,<br>HOSP | FPV, SOC | 780 |
| NCT04324463 | Brazil | 2020-Dec | OUTPAT,<br>HOSP | HCQ + AZM, SOC | 4000 |
| NCT04393038 | Belgium | 2020-Dec | OUTPAT,<br>HOSP | Other, SOC | 1034 |
| CHICTR2000029954 | China | 2020-Feb | OUTPAT,<br>HOSP | SOC, Herbal tea | 300 |
| CHICTR2000030102 | China | 2020-May | OUTPAT,<br>HOSP | Non-invasive respiratory support, SOC | 180 |
| NCT04338828 | United States | 2021-Apr | OUTPAT,<br>HOSP | Nitric Oxide, SOC | 260 |
| NCT04390191 | United States | 2021-Apr | OUTPAT,<br>HOSP | Non-invasive respiratory support, SOC | 200 |
| NCT04517396 | United States | 2021-Aug | OUTPAT,<br>HOSP | BIO101 (MAS activator),SOC | 300 |
| IRCT20190810044500N5 | Iran | 2021-Feb | OUTPAT,<br>HOSP | Colchicine, HCQ + Colchicine | 200 |
| CHICTR2000029867 | China | 2021-Feb | OUTPAT,<br>HOSP | Carrimycin, LPV/r | 520 |
| NCT04394117 | Australia | 2021-Jan | OUTPAT,<br>HOSP | ARBs, SOC | 605 |
| NCT04419025 | United States | 2021-Jan | OUTPAT,<br>HOSP | N-acetyl ccysteine | 200 |
| NCT04500418 | Germany | 2021-Sep | OUTPAT,<br>HOSP | cenicriviroc (CCR2 and CCR5 receptor blocker), SOC | 183 |
| CTRI/2020/06/025763 | India | NA | OUTPAT,<br>HOSP | Alternative therapy | 150 |
| ISRCTN14212905 | United Kingdom | NA | OUTPAT,<br>HOSP | Anticoagulants, Other, SOC | 100 |
| 2020-001498-63 | Belgium | NA | OUTPAT,<br>HOSP | BIO101 (MAS activator) | 465 |
| CHICTR2000029573 | China | NA | OUTPAT,<br>HOSP | cytokine derived gene protein,SOC | 480 |
| NCT04445467 | Australia | NA | OUTPAT,<br>HOSP | FPV, SOC | 190 |
| NCT04336332 | United States | NA | OUTPAT,<br>HOSP | HCQ, HCQ + AZM, SOC | 160 |
| NCT04385095 | United Kingdom | NA | OUTPAT,<br>HOSP | IFN, SOC | 820 |
| ISRCTN14241621 | United Kingdom | NA | OUTPAT,<br>HOSP | IFN, SOC | 220 |
| NCT04354259 | Canada | NA | OUTPAT,<br>HOSP | IFN, SOC | 140 |
| 2020-001860-27 | United Kingdom | NA | OUTPAT,<br>HOSP | multiple drugs, SOC | 250 |
| NCT04377308 | United States | NA | OUTPAT,<br>HOSP | SOC, SSRIs | 2000 |
| 2020-001501-24 | Italy | NA | PREV | HCQ, SOC | 2300 |
| NCT04374942 | Canada | 2020-Aug | PREV | HCQ, SOC | 988 |
| NCT04359537 | Pakistan | 2020-Aug | PREV | HCQ, SOC | 200 |
| CHICTR2000029602 | China | 2020-Aug | PREV | Education + Monitoring + Herbal tea, Psychological therapy | 600 |
| NCT04380701 | Germany | 2020-Aug | PREV | BNT162 RNA Vaccine | 200 |
| NCT04334928 | Spain | 2020-Dec | PREV | HCQ, HCQ + NRTIs, NRTIs, SOC | 4000 |
| NCT04318015 | Mexico | 2020-Dec | PREV | HCQ, SOC | 400 |
| NCT04348435 | United States | 2020-Dec | PREV | SOC, Stem cells | 100 |
| CHICTR2000031944 | China | 2020-Dec | PREV | SOC, Herbal Tea | 2000 |
| NCT04530357 | Kazakhstan | 2020-Dec | PREV | QAZ-Covid Inactivated Vaccine, SOC | 244 |
| NCT04344379 | France | 2020-Jul | PREV | AZM, HCQ, SOC | 900 |
| NCT04349228 | Tunisia | 2020-Jul | PREV | HCQ, SOC | 530 |
| NCT04366180 | Spain | 2020-Jun | PREV | Nutrition, SOC | 314 |
| NCT04320238 | China | 2020-May | PREV | IFN, IFN + Other | 2944 |
| NCT04421391 | United States | 2020-Nov | PREV | Nutrition | 500 |
| NCT04466280 | Iran | 2020-Nov | PREV | HCQ + mucodentol, mucodentol | 180 |
| NCT04352933 | United Kingdom | 2020-Oct | PREV | HCQ, SOC | 1000 |
| NCT04521322 | Argentina | 2020-Oct | PREV | Iota-Carageenan | 400 |
| NCT04353128 | Spain | 2020-Oct | PREV | Melatonin, SOC | 450 |
| NCT04313023 | United States | 2020-Oct | PREV | PUL-042 (toll-like receptor agonists, SOC | 200 |
| IRCT20200401046909N2 | Iran | 2020-Oct | PREV | SOC, Vitamin D3 | 540 |
| NCT04405908 | Australia | 2020-Oct | PREV | SCB019 (Clover), SOC | 150 |
| NCT04495933 | Australia | 2020-Oct | PREV | SOC, M59 clamp Vaccine | 216 |
| NCT04384458 | Brazil | 2020-Sep | PREV | HCQ, SOC | 400 |
| NCT04409327 | United States | 2020-Sep | PREV | Other, SOC | 550 |
| NCT04303507 | Thailand | 2021-Apr | PREV | HCQ, SOC | 40000 |
| NCT04349371 | United States | 2021-Apr | PREV | HCQ, SOC | 350 |
| NCT04312243 | United States | 2021-Apr | PREV | Non-invasive respiratory support, SOC | 470 |
| NCT04353037 | United States | 2021-Apr | PREV | HCQ, SOC | 850 |
| NCT04435808 | United States | 2021-Apr | PREV | HCQ, SOC | 350 |
| NCT04321928 | Hungary | 2021-Apr | PREV | Education, Psychological therapy | 7576 |
| NCT04368728 | United States | 2021-Apr | PREV | SOC, Vaccine Pfizer Biontech mRNA Vaccine | 29481 |
| NCT04333732 | United States | 2021-Aug | PREV | HCQ, SOC | 30000 |
| NCT04449276 | Belgium | 2021-Aug | PREV | SOC, CVnCov Vaccine | 168 |
| NCT04461379 | Mexico | 2021-Jan | PREV | SOC, Vaccine | 908 |
| CHICTR2000034780 | China | 2021-Jul | PREV | SOC, Vaccine | 15000 |
| IRCT20200411047019N1 | Iran | 2021-Jun | PREV | SOC, BCG | 500 |
| NCT04327206 | Australia | 2021-Jun | PREV | Vaccine | 10078 |
| NCT04471519 | India | 2021-Jun | PREV | SOC, Vaccine | 1125 |
| NCT04363450 | United States | 2021-Mar | PREV | HCQ, SOC | 1700 |
| NCT04448119 | Canada | 2021-Mar | PREV | FPV, SOC | 760 |
| NCT04364022 | Switzerland | 2021-Mar | PREV | HCQ, LPV/r | 300 |
| NCT04318444 | United States | 2021-Mar | PREV | HCQ, SOC | 1600 |
| NCT04321174 | Canada | 2021-Mar | PREV | LPV/r, SOC | 1220 |
| NCT04348370 | United States | 2021-May | PREV | SOC, Vaccine | 1800 |
| NCT04364802 | United States | 2021-May | PREV | Hygiene, SOC | 250 |
| NCT04414267 | Greece | 2021-May | PREV | SOC, Vaccine | 900 |
| NCT04530396 | Russia | 2021-May | PREV | SOC, Vaccine Russian Gamelaya Vaccine | 40000 |
| NCT04519437 | United States | 2021-Oct | PREV | mAb, SOC | 940 |
| NCT04456595 | Brazil | 2021-Sep | PREV | SOC, Vaccine | 8870 |
| NCT04536051 | Brazil | 2021-Sep | PREV | Vaccine | 5000 |
| NCT04537949 | Germany | 2021-Sep | PREV | BNT162 RNA Vaccine | 120 |
| NCT04466085 | China | 2021-Sep | PREV | SOC, Vaccine | 900 |
| NCT04372017 | United States | 2022-Apr | PREV | HCQ, Vitamins | 1739 |
| NCT04323800 | United States | 2022-Dec | PREV | Plasma based therapy, SOC | 500 |
| NCT04447781 | South Korea | 2022-Feb | PREV | Device, INO4800 + Device | 160 |
| NCT04299724 | China | 2023-Jul | PREV | aAPC Vaccine | 100 |
| CTRI/2020/05/025275 | India | NA | PREV | Alternative therapy, SOC | 200 |
| NCT04359680 | United States | NA | PREV | Antivirals, SOC | 800 |
| ACTRN12620000417987 | Australia | NA | PREV | HCQ | 680 |
| NCT04340349 | Mexico | NA | PREV | HCQ, bromhexine, SOC | 140 |
| CTRI/2020/05/025067 | India | NA | PREV | HCQ, SOC | 6950 |
| NCT04341441 | United States | NA | PREV | HCQ, SOC | 3000 |
| ACTRN12620000501943 | Australia | NA | PREV | HCQ, SOC | 2250 |
| NCT04334148 | United States | NA | PREV | HCQ, SOC | 2000 |
| 2020-001987-28 | Italy | NA | PREV | HCQ, SOC | 1000 |
| NCT04349228 | Tunisia | NA | PREV | HCQ, SOC | 530 |
| NCT04496245 | Australia | NA | PREV | Immunomodulators, SOC | 1000 |
| NCT04452773 | Spain | NA | PREV | Manremyc food supplement | 315 |
| NCT04408183 | United States | NA | PREV | quinine topical nasal spary | 225 |
| NCT04379336 | South Africa | NA | PREV | SOC, BCG | 500 |
| ISRCTN50212645 | United Kingdom | NA | PREV | Other | 2000 |
| CTRI/2020/05/025429 | India | NA | PREV | Alternative therapy, SOC | 5000 |
| NCT04343248 | United States | NA | PREV | Antivirals, SOC | 800 |
| SLCTR/2020/011 | Sri Lanka | NA | PREV | HCQ, SOC | 400 |
| CTRI/2020/06/025928 | India | NA | PREV | Hygiene | 204 |
| NCT04344600 | United States | NA | PREV | IFN, SOC | 164 |
| NCT04399252 | United States | NA | PREV | Nutrition, SOC | 1000 |
| CTRI/2020/05/025277 | India | NA | PREV | Other, SOC | 4000 |
| CHICTR2000032459 | China | NA | PREV | SOC, Inactivated SARS-Cov-2 (Vero cell) Vaccine | 288 |
| NCT04475302 | India | NA | PREV | SOC, Vaccine | 2175 |
| NCT04368988 | United States | NA | PREV | SOC, Vaccine | 1631 |
| NCT04369794 | Brazil | NA | PREV | SOC, Vaccine | 1000 |
| NCT04470427 | United States | NA | PREV | SOC, Vaccine mRNA1273 | 30000 |
| CTRI/2020/06/025769 | India | NA | PREV | Alternative therapy, SOC | 40000 |
| CTRI/2020/05/025484 | India | NA | PREV | Alternative therapy, SOC | 5000 |
| CTRI/2020/05/025491 | India | NA | PREV | Alternative therapy, SOC | 1000 |
| CTRI/2020/07/026560 | India | NA | PREV | Alternative therapy, SOC | 200 |
| IRCT20130306012728N8 | Iran | NA | PREV | HQC, SOC | 500 |
| NCT04387409 | Germany | NA | PREV | SOC, Vaccine | 1200 |
| NCT04384549 | France | NA | PREV | SOC, Vaccine | 1120 |
| NCT04381988 | United States | NA | PREV | HQC + Radiation therapy, Radiation therapy | 132 |
| 2020-002287-31 | Spain | NA | PREV | HQC, SOC | 1930 |
| ISRCTN17072692 | United Kingdom | NA | PREV | LNP-nCoVsaRNA Vaccine | 320 |
| NCT04357028 | Egypt | NA | PREV | measles vaccine, SOC | 200 |
| NCT04445389 | South Korea | NA | PREV | SOC, GX-19 DNA Vaccine | 210 |
| NCT04470609 | China | NA | PREV | SOC, SARS-Cov-2 inactivated vaccine | 471 |
| NCT04439045 | Canada | NA | PREV | SOC, Vaccine | 3626 |
| NCT04445428 | Guinea-Bissau | NA | PREV | SOC, Vaccine | 3400 |
| ISRCTN89951424 | Brazil | NA | PREV | SOC, Vaccine | 2000 |
| NL8547 | Netherlands | NA | PREV | SOC, Vaccine | 1600 |
| NCT04442048 | Canada | NA | PREV | SOC, Vaccine | 1500 |
| ISRCTN15281137 | United Kingdom | NA | PREV | SOC, Vaccine | 1112 |
| CTRI/2020/07/026352 | India | NA | PREV | SOC, Vaccine | 1048 |
| NCT04436276 | United States | NA | PREV | SOC, Vaccine | 1045 |
| NCT04412538 | China | NA | PREV | SOC, Vaccine | 942 |
| RPCEC00000314 | Cuba | NA | PREV | SOC, Vaccine Finlay Vaccine Institute | 115000 |
| ISRCTN90906759 | United Kingdom | NA | PREV | Vaccine | 12330 |
| CTRI/2020/06/025861 | India | NA | PREV | Alternative therapy, SOC | 500 |
| NCT04498442 | United States | NA | PREV | Exercise, Other, Respiratory support | 30000 |
| CHICTR2000030937 | China | 2020-Aug | TAIL | SOC, Herbal tea | 144 |
| CHICTR2000030720 | China | 2020-Aug | TAIL | SOC, Herbal tea | 120 |
| NCT04368845 | Greece | 2020-Jul | TAIL | Non-invasive respiratory support, SOC | 100 |
| CHICTR2000030987 | China | 2020-Jun | TAIL | FPV, FPV + HCQ, SOC | 150 |
| NCT04333589 | China | 2020-Jun | TAIL | FPV, SOC | 210 |
| NCT04394169 | Spain | 2020-Sep | TAIL | Rehabilitation, SOC | 102 |
| CHICTR2000029789 | China | 2021-Jun | TAIL | SOC, Herbal tea | 100 |
| CHICTR2000030920 | China | 2021-May | TAIL | SOC, Herbal tea | 100 |
| NCT04351880 | United States | NA | TAIL | Meal planning | 640 |
| NCT04279197 | China | NA | TAIL | SOC, Herbal tea | 160 |
| CHICTR2000029578 | China | 2020-Apr | UNCLEAR | Alternate therapy | 1000 |
| NCT04365309 | China | 2020-Apr | UNCLEAR | Antihypertensive, SOC | 128 |
| NCT04319900 | China | 2020-Apr | UNCLEAR | FPV, HCQ + FPV, SOC | 150 |
| CHICTR2000031204 | China | 2020-Apr | UNCLEAR | HCQ, SOC | 300 |
| CHICTR2000030923 | China | 2020-Apr | UNCLEAR | SOC, Herbal tea | 400 |
| CHICTR2000029517 | China | 2020-Apr | UNCLEAR | SOC, Herbal tea | 100 |
| NCT04326790 | Gibraltar | 2020-Aug | UNCLEAR | Colchicine, SOC | 180 |
| NCT04527562 | Bangladesh | 2020-Dec | UNCLEAR | Colchicine, SOC | 300 |
| CHICTR2000032915 | China | 2020-Dec | UNCLEAR | HCQ, AZM, SOC | 240 |
| CHICTR2000030219 | China | 2020-Dec | UNCLEAR | Herbal tea | 120 |
| NCT04447235 | Brazil | 2020-Dec | UNCLEAR | Ivermectin + ARBs, SOC | 176 |
| NCT04323345 | Egypt | 2020-Dec | UNCLEAR | Nutrition, SOC | 1000 |
| CHICTR2000033320 | China | 2020-Dec | UNCLEAR | SOC, Herbal tea | 300 |
| CHICTR2000030288 | China | 2020-Dec | UNCLEAR | SOC, Herbal tea | 204 |
| CHICTR2000032098 | China | 2020-Jul | UNCLEAR | Herbal tea | 300 |
| CHICTR2000030480 | China | 2020-Jul | UNCLEAR | IFN, SOC | 328 |
| NCT04425707 | Egypt | 2020-Jul | UNCLEAR | Ivermectin, SOC | 100 |
| CHICTR2000032399 | China | 2020-Jul | UNCLEAR | SOC, Herbal tea | 200 |
| NCT04334265 | China | 2020-Jun | UNCLEAR | Alternative therapy, SOC | 750 |
| NCT04392141 | Iran | 2020-Jun | UNCLEAR | Colchicine + Alternative therapy, SOC | 200 |
| CHICTR2000032237 | China | 2020-Jun | UNCLEAR | SOC, Herbal tea | 200 |
| CHICTR2000030034 | China | 2020-Jun | UNCLEAR | SOC, Herbal tea | 132 |
| CHICTR2000032400 | China | 2020-Jun | UNCLEAR | SOC, Herbal tea | 120 |
| CHICTR2000030490 | China | 2020-Mar | UNCLEAR | Herbal tea | 100 |
| CHICTR2000030535 | China | 2020-Mar | UNCLEAR | LPV/r + IFN, LPV/r + IFN + Other | 100 |
| CHICTR2000030188 | China | 2020-Mar | UNCLEAR | SOC, Herbal tea | 120 |
| CHICTR2000030810 | China | 2020-Mar | UNCLEAR | SOC, Herbal tea | 100 |
| CHICTR2000029550 | China | 2020-May | UNCLEAR | Herbal tea | 300 |
| CHICTR2000029558 | China | 2020-May | UNCLEAR | Herbal tea | 200 |
| CHICTR2000029549 | China | 2020-May | UNCLEAR | SOC, Herbal tea | 400 |
| CHICTR2000029778 | China | 2020-May | UNCLEAR | SOC, Herbal tea | 600 |
| NCT04405843 | Colombia | 2020-Nov | UNCLEAR | Ivermectin, SOC | 400 |
| NCT04355936 | Argentina | 2020-Oct | UNCLEAR | Antihypertensive, SOC | 400 |
| NCT04312997 | United States | 2020-Oct | UNCLEAR | PUL-042 (toll-like receptor agonists, SOC | 100 |
| CHICTR2000029790 | China | 2020-Oct | UNCLEAR | SOC, Herbal tea | 120 |
| NCT04353336 | Egypt | 2020-Sep | UNCLEAR | HCQ | 200 |
| NCT04325893 | France | 2020-Sep | UNCLEAR | HCQ, SOC | 1300 |
| NCT04429711 | Israel | 2020-Sep | UNCLEAR | Ivermectin, SOC | 100 |
| NCT04463264 | Argentina | 2020-Sep | UNCLEAR | Nitazoxanide, SOC | 135 |
| NCT04338698 | Pakistan | 2020-Sep | UNCLEAR | Oseltamavir vs Azithromycin vs HCQ | 500 |
| NCT04449588 | China | 2021-Aug | UNCLEAR | BDB001 (toll like receptor agonist), SOC | 368 |
| CHICTR2000030117 | China | 2021-Dec | UNCLEAR | LPV/r + IFN, Herbal tea | 348 |
| CHICTR2000029935 | China | 2021-Feb | UNCLEAR | HCQ | 100 |
| CHICTR2000029939 | China | 2021-Feb | UNCLEAR | HCQ, SOC | 100 |
| NCT04421508 | United States | 2021-Feb | UNCLEAR | Non-invasive respiratory support, SOC | 500 |
| CHICTR2000029605 | China | 2021-Feb | UNCLEAR | SOC, Herbal tea | 400 |
| CHICTR2000029747 | China | 2021-Feb | UNCLEAR | SOC, Herbal tea | 200 |
| IRCT20110726007117N11 | Iran | 2021-Jan | UNCLEAR | SOC, Vitamin D | 210 |
| NCT04334512 | United States | 2021-Jun | UNCLEAR | HCQ,AZM, Vitamins, Zinc | 600 |
| NCT04344041 | France | 2021-May | UNCLEAR | ,Vitamin D, SOC | 260 |
| CHICTR2000032919 | China | 2021-May | UNCLEAR | SOC, Herbal tea | 200 |
| NCT04395768 | Australia | 2021-Sep | UNCLEAR | HCQ + AZM + Zinc + Vitamins | 200 |
| CHICTR2000029756 | China | 2022-Feb | UNCLEAR | IFN, Herbal tea | 238 |
| CHICTR2000029976 | China | 2022-Feb | UNCLEAR | Psychological therapy, SOC | 160 |
| CTRI/2020/06/026221 | India | NA | UNCLEAR | Alternative therapy, HCQ | 100 |
| CTRI/2020/06/025557 | India | NA | UNCLEAR | Alternative therapy, SOC | 420 |
| CTRI/2020/07/026468 | India | NA | UNCLEAR | Alternative therapy, SOC | 140 |
| NCT04351347 | Egypt | NA | UNCLEAR | Antivirals, HCQ, Ivermectin | 300 |
| CHICTR2000032769 | China | NA | UNCLEAR | Azuvidine, SOC | 342 |
| NCT04471766 | Guinea-Bissau | NA | UNCLEAR | Cloth face mask, SOC | 66000 |
| CTRI/2020/05/025320 | India | NA | UNCLEAR | Exercise + Nutrition + Hygiene + Alternative therapy + Other, SOC | 658 |
| CTRI/2020/06/025575 | India | NA | UNCLEAR | HCQ + ribavirin, LPV/r + ribavirin | 175 |
| NCT04374019 | United States | NA | UNCLEAR | HCQ, HCQ + AZM, HCQ + Ivermectin, Other | 240 |
| NCT04447534 | Egypt | NA | UNCLEAR | HCQ, HCQ + Zinc | 200 |
| NCT04403555 | Egypt | NA | UNCLEAR | HCQ, Ivermectin + Docycline | 200 |
| NCT04410562 | Spain | NA | UNCLEAR | HCQ, SOC | 714 |
| NCT04352959 | France | NA | UNCLEAR | Hygiene, SOC | 178 |
| RPCEC00000307 | Cuba | NA | UNCLEAR | IFN | 120 |
| CHICTR2000029638 | China | NA | UNCLEAR | IFN | 100 |
| CHICTR2000030262 | China | NA | UNCLEAR | IFN, SOC | 140 |
| NCT04255017 | China | NA | UNCLEAR | LPV/r, Oseltamivir, SOC, Umifenovir | 400 |
| NCT04495816 | United States | NA | UNCLEAR | Nutrition, SOC | 126 |
| 2020-001736-95 | United Kingdom | NA | UNCLEAR | Other, SOC | 825 |
| NCT04407390 | Denmark | NA | UNCLEAR | SOC, nicotamid riboside | 100 |
| NCT04510207 | Bahrain | NA | UNCLEAR | SOC, Vaccine Wuhan Institute of Biological Products Vero cell | 45000 |
| JPRN-JRCTS061200002 | Japan | NA | UNCLEAR | teprenone, SOC | 100 |
| NCT04254874 | China | NA | UNCLEAR | Umifenovir, Umifenovir + IFN | 100 |
| NCT04407286 | United States | NA | UNCLEAR | Vitamin D | 100 |

| ID | Country | Completion | Patient setting | Treatment |
| --- | --- | --- | --- | --- |
| NCT04575038 | United States | 2021-Jan | Outpatient | DHODH inhibitor brequinar. SOC |
| NCT04564716 | Belarus | 2021-Mar | Healthy Unexposed | SOC, GAM-Covid Vaccine |
| NCT04610567 | Brazil | 2021-Mar | Hospital | Methotrexate, SOC |
| NCT04509999 | United States | 2022-Sep | Outpatient | Other, SOC |
| NCT04432324 | Spain | 2020-Dec | Hospital | iViG |
| NCT04377568 | Canada | 2021-Dec | Hospital | Plasma based therapy, SOC |
| NCT04392713 | Pakistan | NA | NA | Ivermectin, SOC |
| NCT04466683 | United States | NA | Hospital | Radiation therapy, SOC |
| NCT04492228 | Italy | NA | Hospital | Nutrition, SOC |
| CTRI/2020/08/026957 | India | NA | Hospital | Alternative therapy, SOC |
| PACTR202006760881890 | Nigeria | NA | Hospital | Plasma based therapy, SOC |
| RPCEC00000309 | Mexico | NA | Outpatient, Hospital, ICU | activated sodium chloride inhalation |
| NCT04420676 | Austria | NA | Outpatient | Nutrition, SOC |
| NCT04549922 | Brazil | 2021-Feb | Unclear | Antisense therapy, SOC |
| NCT04587219 | Russia | 2020-Dec | NA | GAM-Covid vaccine |
| NCT04510493 | Switzerland | 2023-Sep | Hospital | mAb, SOC |
| JPRN-JRCT2071200023 | Japan | NA | Unclear | viracept, SOC |
| CTRI/2020/06/026001 | India | NA | Hospital | Ivermectin, SOC |
| CTRI/2020/08/027106 | India | NA | Hospital | Alternative therapy, SOC |
| NCT04596085 | India | 2020-Nov | Outpatient | Nutrition, SOC |
| NCT04472494 | United States | 2021-Jan | Hospital | abatacept |
| NCT04410328 | United States | 2021-Mar | Hospital | anticoagulant, aggrenox |
| NCT04516759 | United Kingdom | 2021-Mar | Hospital | glucokinase activator |
| NCT04551911 | United States | 2021-Feb | Unclear | SOC, Vitamins rayaldee |
| NCT04604327 | Spain | 2021-May | Hospital | Anticoagulants |
| NCT04576728 | Spain | 2021-Mar | Hospital | SOC, Trimodulin |
| NCT04523181 | United States | 2020-Dec | Hospital | antroquinolol |
| NCT04511650 | United States | 2021-Feb | Hospital | Razuprotafib, SOC |
| NCT04428021 | Italy | NA | Hospital | Plasma based therapy, SOC |
| PER-013-20 | Peru | NA | Hospital, ICU | Plasma based therapy, SOC |
| NCT04578158 | Pakistan | 2021-Apr | Hospital | Alternative therapy, SOC |
| NCT04583956 | United States | 2021-Jan | Hospital | Remdesivir, Remdesivir + mAb |
| NCT04558021 | Turkey | 2021-Jan | NA | Niclosamide |
| 2020-001329-30 | China | NA | ICU | Non-invasive respiratory support |
| ACTRN12620000557932 | Australia | NA | Hospital | HCQ + Zinc + Vitamins + AZT |
| CTRI/2020/06/025768 | India | NA | Unclear | Alternative therapy |
| NCT04622865 | France | 2021-Jun | Hospital | Alternative therapy + Other, HCQ |
| NCT04357457 | France | NA | Hospital | Respiratory stimulant, SOC |
| NCT04499677 | United Kingdom | NA | Outpatient | FPV, LPV/r, LPV/r + FPV, SOC |
| NCT04523246 | United States | 2021-Jul | Nursing home residents | SOC, shingrix Vaccine |
| NCT04568031 | Japan | 2021-Sep | NA | Vaccine ChAdOx1 Vector Vaccine (nn-replicating) |
| NCT04498247 | United States | 2022-Apr | NA | V591 Merck Vaccine |
| NCT04583592 | United States | 2020-Dec | Outpatient | Camostat, SOC |
| 2020-002713-17 | Mexico | NA | Hospital | Immunomodulators, mAb, SOC |
| NCT04370236 | United States | 2021-Feb | Hospital | INBO3, checkpoint inhibitor |
| NCT04455815 | United Kingdom | 2021-Aug | Outpatient | Camostat, SOC |
| NCT04530435 | Denmark | 2021-Apr | Outpatient | Respiratory support, SOC |
| NCT04535674 | Spain | NA | Hospital | Other, SOC |
| 2020-001662-11 | France | NA | Hospital | JAKi, SOC |
| NCT04603924 | United States | 2021-Jun | Hospital | Other, SOC Niclosamide |
| NCT04494646 | United States | 2021-Jan | Hospital | SOC, Triterpenes |
| NCT04435795 | Canada | 2021-Feb | Outpatient | Corticosteroids, SOC |
| NCT04581954 | United Kingdom | 2021-Jun | Hospital | Immunomodulators, JAKi, SOC |
| NCT04377750 | Israel | 2020-Apr | ICU | SOC, TCZ |
| NCT04321096 | Denmark | 2020-Dec | Hospital | Other, SOC |
| NCT04504032 | United States | 2020-Dec | Outpatient | Anticoagulants, SOC |
| CHICTR2000039000 | China | 2020-Dec | NA | SOC, Inactivated SARS Cov 2 vaccine |
| NCT04335084 | United States | 2021-Jun | Healthcare Workers | HCQ + Vitamins + Zinc, SOC |
| NCT04471636 | Germany | NA | Outpatient | Device + Other, SOC |
| NCT04589949 | Netherlands | 2022-Nov | Outpatient | Plasma-based therapy |
| NCT04376684 | United States | 2020-Dec | Hospital | mAb, SOC |
| CTRI/2020/07/026791 | India | NA | Hospital | Antihypertensive, SOC, Statins |
| NCT04350593 | United States | 2020-Oct | Hospital | SGLT2, SOC |
| NCT04504734 | United States | 2021-Feb | Outpatient | Antirheumatic, SOC bucillamine |
| ACTRN12620000473965 | Australia | NA | Healthcare Workers | Immunomodulators, SOC |
| NCT04492475 | United States | 2023-Nov | Hospital | Remdesivir, Remdesivir + IFN |
| 2020-001072-15 | United Kingdom | NA | Healthy Unexposed | Vaccine |
| NCT04359095 | Colombia | 2020-Nov | Hospital | AZT, HCQ, LPV/r, SOC |
| NCT04501952 | United States | 2020-Dec | Outpatient | Remdesivir, SOC |
| NCT04575584 | United States | 2021-Apr | NA | Antivirals molnupiravir, SOC |
| NCT04545060 | United States | 2021-Jan | Outpatient | Monoclonal antibody, SOC |
| NCT04606563 | United States | 2021-Jun | NA | ARBs, SOC |
| NCT04575597 | United States | 2021-Apr | Outpatient | Antivirals, SOC |
| CTRI/2020/08/027170 | India | NA | Healthy Unexposed | SOC, Vaccine Covishield |
| NCT04444674 | South Africa | 2020-Oct | NA | SOC, Vaccine ChAdOx1 nCoV-19) |
| NCT04593940 | United States | 2021-Feb | Hospital | Remdesivir + Costimulation blocker, Remdesivir + mAb, Remdesivir + Other |
| NCT04497987 | United States | NA | Nursing home residents | LY COV 555 monoclonal ab |
| ACTRN12620000445976 | New Zealand | NA | Hospital | HCQ, LPV/r, LPV/r + HCQ |
| NCT04366960 | Italy | 2020-Aug | Hospital | Anticoagulants |
| CHICTR2000030936 | China | 2020-May | Outpatient, Hospital | SOC, TCM |
| NCT04508023 | United States | 2021-Mar | NA | Anticoagulants, SOC |
| NCT04533399 | South Africa | NA | Healthy Unexposed | SOC, Vaccine,Novavax vaccine |
| NCT04498273 | United States | 2021-Sep | Outpatient | Anticoagulants, Antihypertensive, SOC |
| NCT04583995 | United Kingdom | NA | NA | SOC, SOC + Vaccine,,SARS-CoV-2 rS with Matrix-M1 adjuvant,Novavax vaccine |
| 2020-001228-32 | United Kingdom | NA | Healthy Unexposed | SOC, chadOxV vaccine astra zeneca |
| NCT04582344 | Turkey | 2021-Feb | NA | SOC, Vaccine.coronavac vaccine |
| 2020-001441-39 | United Kingdom | NA | Healthcare Workers | HCQ, SOC |
| ISRCTN10207947 | Indonesia | NA | Healthcare Workers | HCQ, SOC |
| NCT04516746 | United States | 2020-Dec | Unclear | SOC, Vaccine,chadOxV vaccine astra zeneca |
| NCT04505722 | United States | NA | Unclear | SOC, Vaccine,JANSEN AD26.Cov2.s |

**SUPPLEMENTARY TABLE 2:**
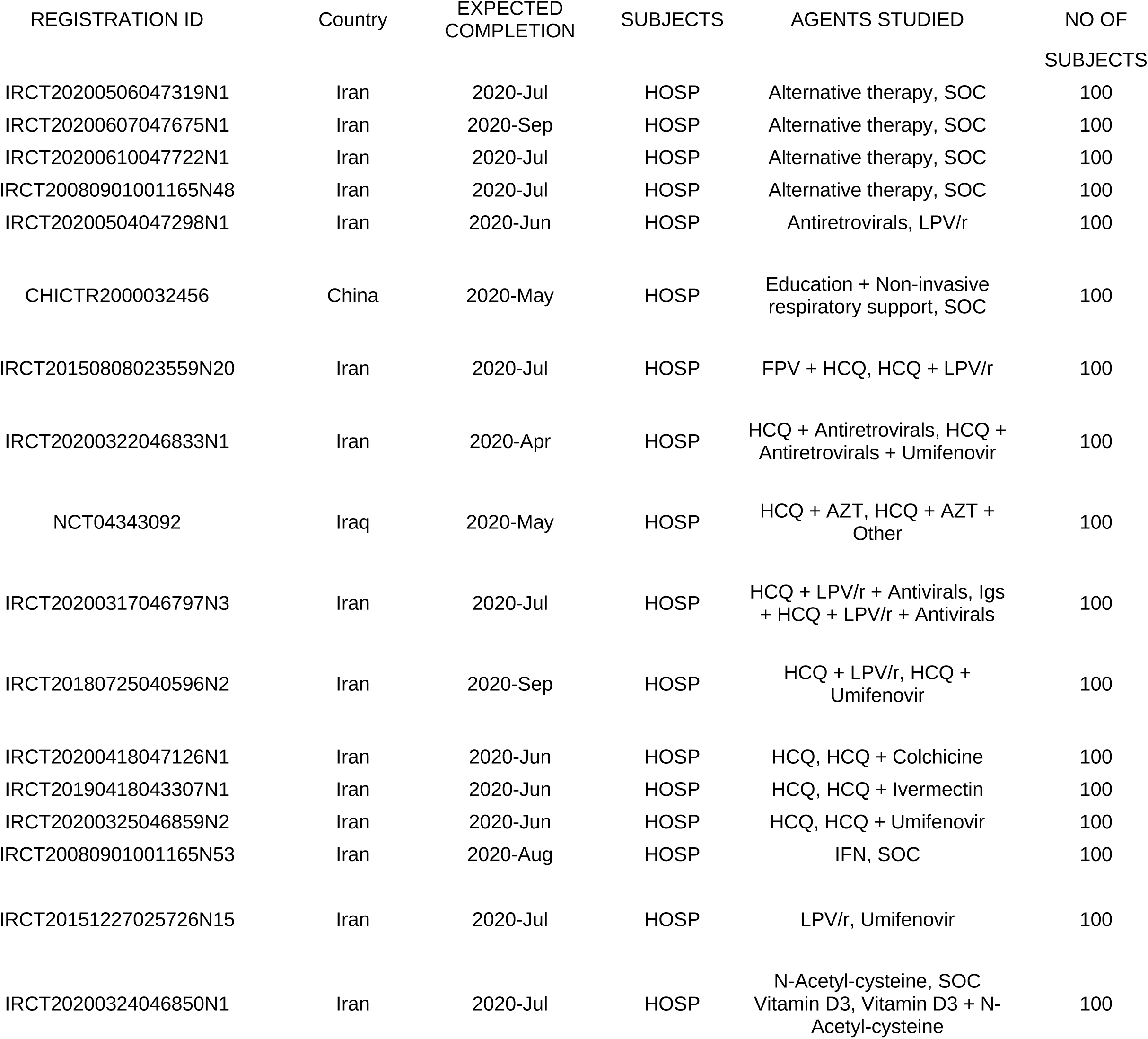
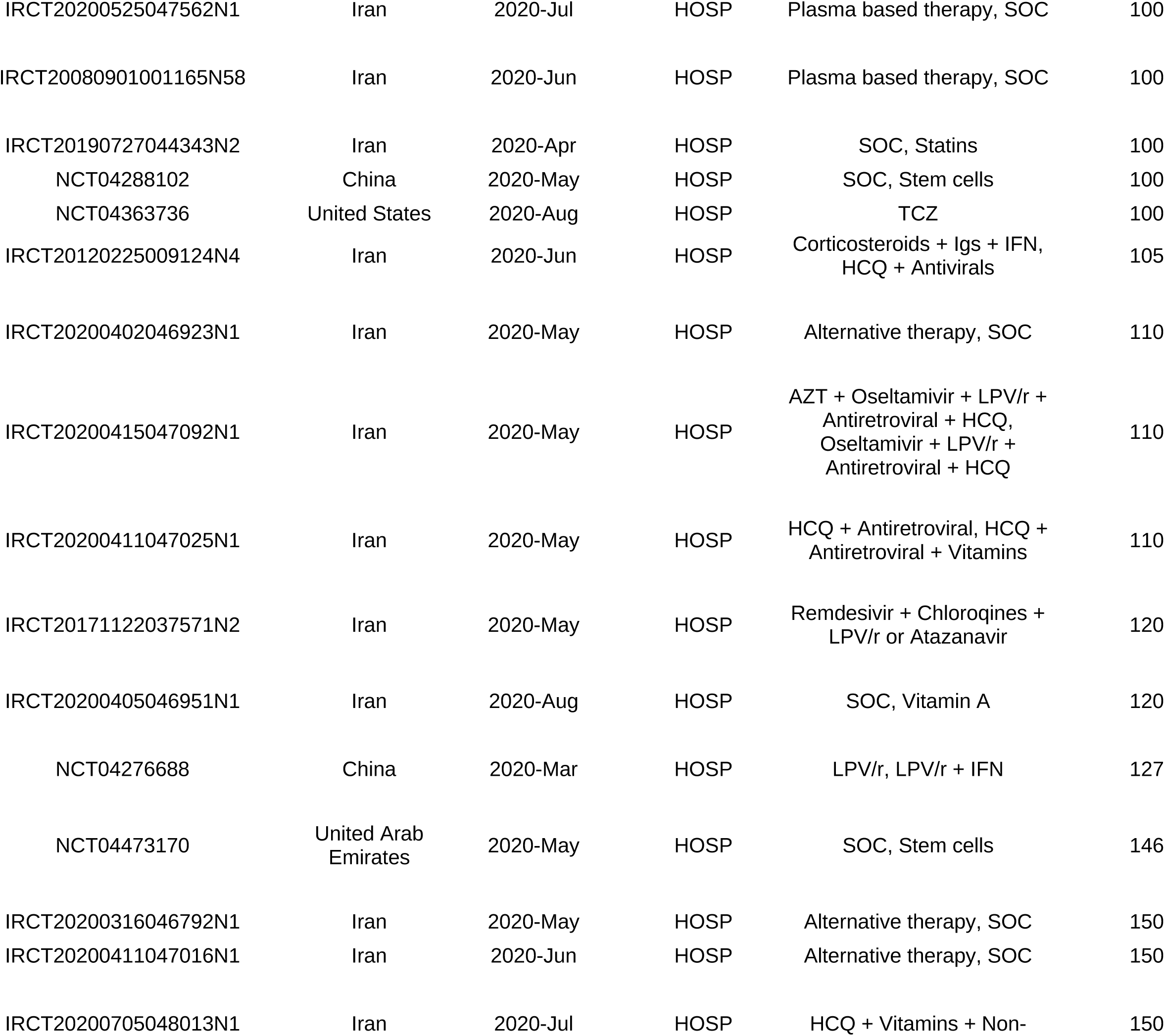
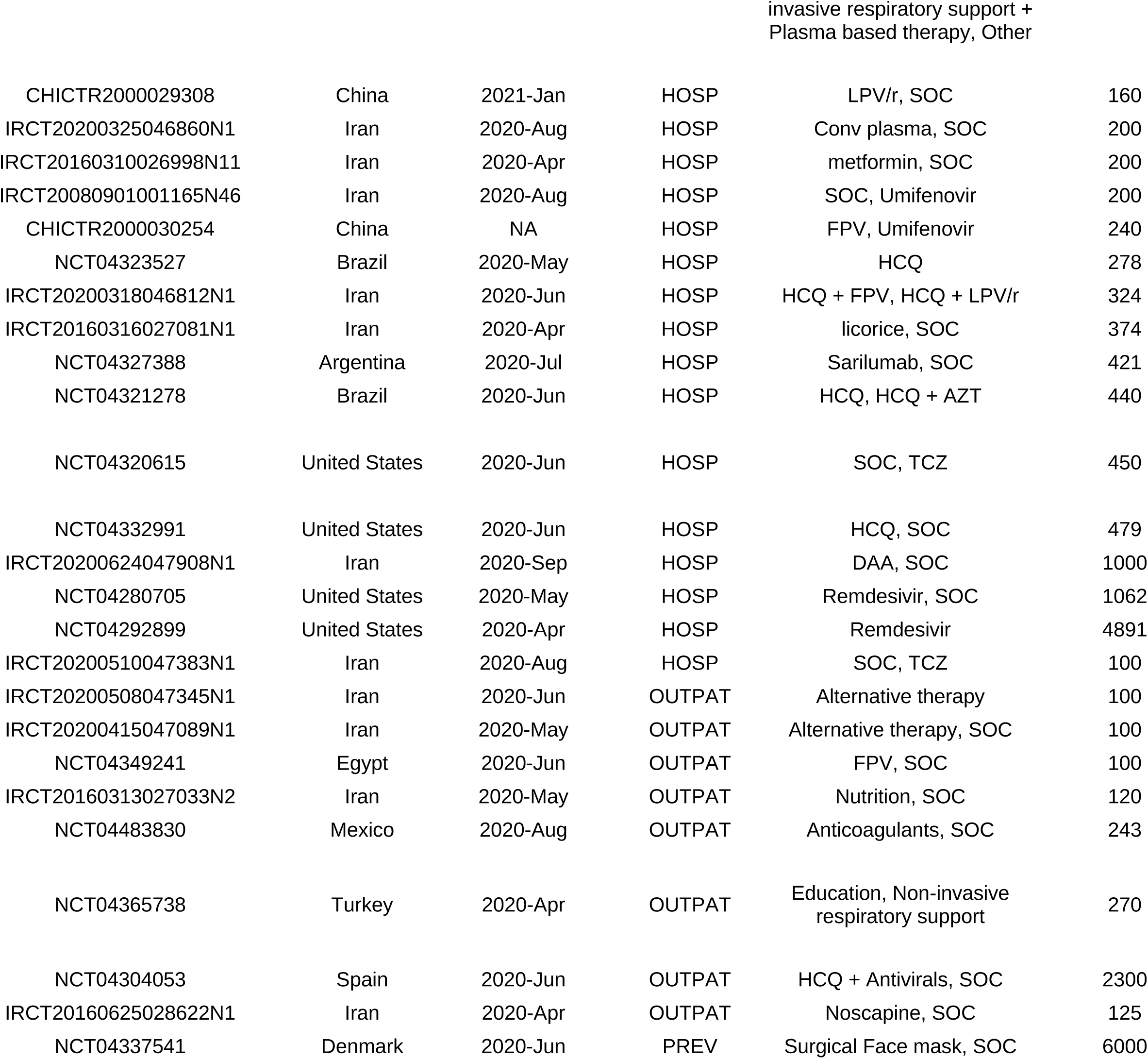
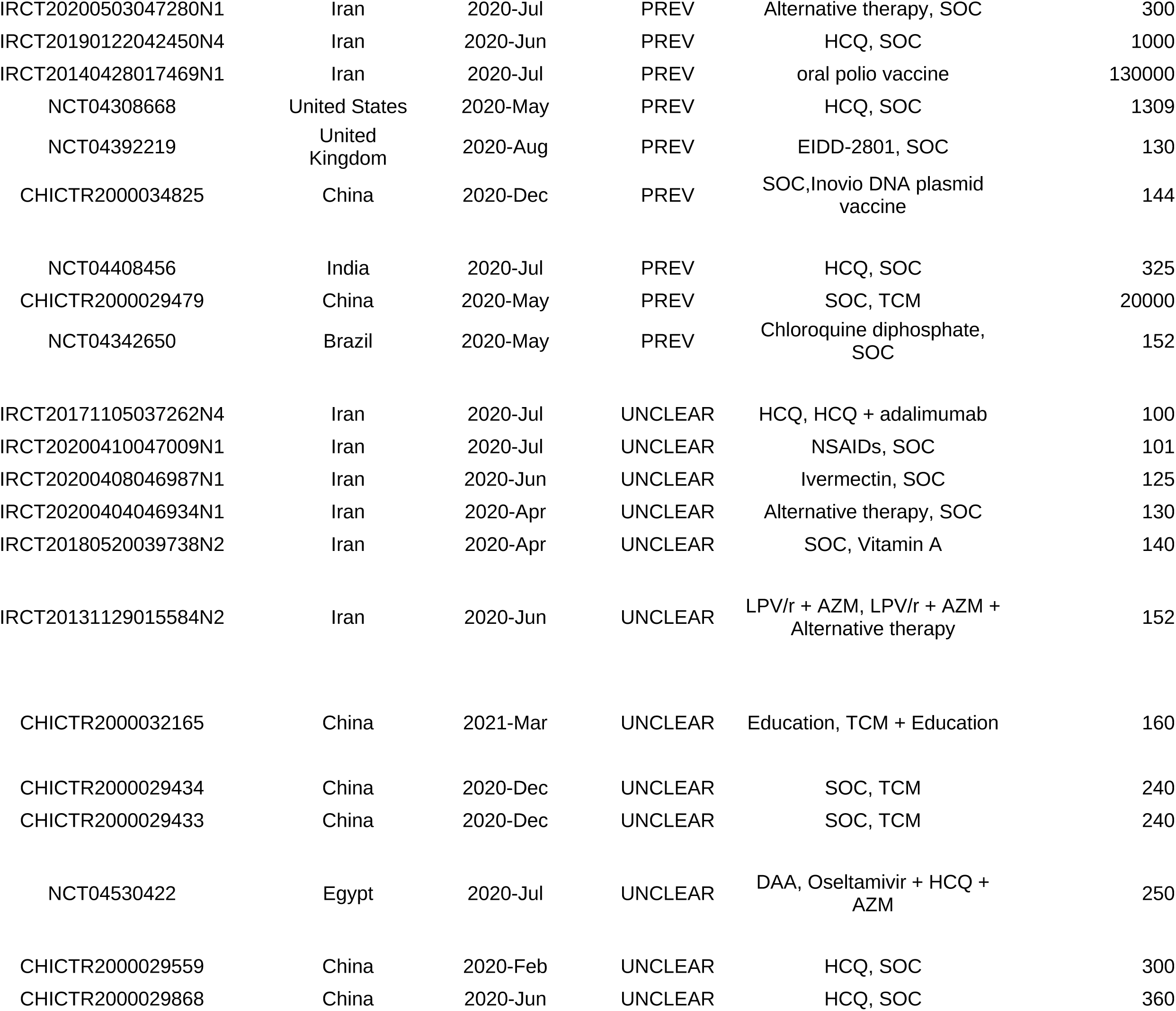
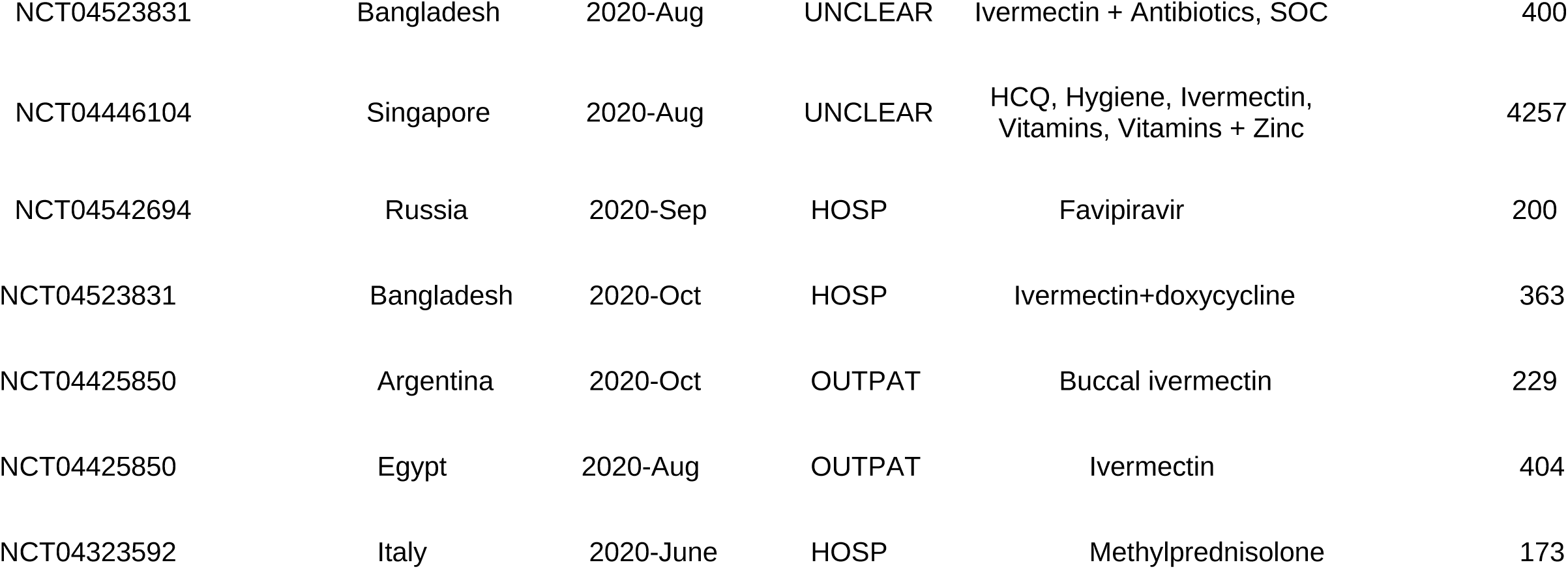
TRIALS COMPLETED AS OF December 27, 2020. The registration ID for the respective regulatory body is given along with the country of registration and the date of completed enrolment. The setting for subjects in each trial: HOSP: Hospitalized; PREV: Healthy or exposed volunteers; OUTPAT: Infected patients, not hospitalized; UNCLEAR: Including multiple categories. SOC: Standard of Care.; FPV: Favipiravir; LPV/R: lopanovir/ritonavir; HCQ: hydroxychloroquine; AZM: azithromycin; TCM: A Chinese herbal tea; NSAID: Non-steroidal anti-inflammatory drug; Conv Plasma: Convalescent plasma

